# COVID-19 vaccine coverage targets to inform reopening plans in a low incidence setting

**DOI:** 10.1101/2022.12.04.22282996

**Authors:** Eamon Conway, Camelia Walker, Chris Baker, Michael Lydeamore, Gerard E. Ryan, Trish Campbell, Joel C. Miller, Max Yeung, Greg Kabashima, James Wood, Nic Rebuli, James M. McCaw, Jodie McVernon, Nick Golding, David J. Price, Freya M. Shearer

## Abstract

Since the emergence of SARS-CoV-2 in 2019 through to mid-2021, much of the Australian population lived in a COVID-19 free environment. This followed the broadly successful implementation of a strong suppression strategy, including international border closures. With the availability of COVID-19 vaccines in early 2021, the national government sought to transition from a state of minimal incidence and strong suppression activities to one of high vaccine coverage and reduced restrictions but with still-manageable transmission. This transition is articulated in the national “re-opening” plan released in July 2021. Here we report on the dynamic modelling study that directly informed policies within the national re-opening plan including the identification of priority age groups for vaccination, target vaccine coverage thresholds and the anticipated requirements for continued public health measures — assuming circulation of the Delta SARS-CoV-2 variant. Our findings demonstrated that adult vaccine coverage needed to be at least 70% to minimise public health and clinical impacts following the establishment of community transmission. They also supported the need for continued application of test-trace-isolate-quarantine and social measures during the vaccine roll-out phase and beyond.

## INTRODUCTION

In early 2020, Australia adopted a strong suppression strategy in response to the COVID-19 pandemic, aiming for no community transmission of the SARS-CoV-2 virus [1]. As a result of a broadly successful implementation of this strategy, which included international and internal travel restrictions, much of the population lived in a COVID-19 free environment up until late 2021. Nonetheless, sporadic outbreaks and a number of major (but geographically isolated) waves of infection occurred, most notably the ‘second wave’ in the state of Victoria June–October 2020 [2] and the Delta ‘third wave’ seeded into New South Wales in June 2021 [3], which quickly spread into neighbouring jurisdictions.

Like elsewhere, the availability of highly effective vaccines for COVID-19 from early 2021 provided a new opportunity to protect the population and reduce harms related to SARS-CoV-2 [4]. However, the transition from a state of minimal (daily and cumulative) incidence and low vaccine coverage to one of high vaccine coverage and established but manageable transmission presented unique challenges compared to vaccination roll-out in high incidence and high pre-existing immunity settings.

In much of Europe and North America, where COVID-19 has circulated widely since its emergence and both daily and cumulative incidence were high at the time of vaccine availability, vaccination has provided a very clear, although challenging, ‘re-opening’ pathway. As vaccine campaigns were initiated, with ancestral and Alpha variants in circulation, increasing vaccine coverage reduced the ability of the virus to spread and assisted in bringing case incidence down [5]. While the emergence of the Delta variant led to a resurgence in cases, there was clear evidence that vaccination played an important role in ‘decoupling’ clinical case loads and deaths from mild infections [6].

In contrast, for countries such as Australia, where pre-vaccination cumulative incidence was very low and the population remained largely susceptible, the relationship between vaccination, incidence and the public perception of COVID-19 was markedly different. Any plan to ‘re-open’ society and transition away from a strong suppression strategy would necessarily lead to an *increase* in cases, morbidity and mortality, with clear challenges for communication and decision making. The targets in the National Plan were agreed by all the States and Territories at National Cabinet on 6 August 2021. The “National Plan to transition Australia’s National COVID-19 Response” (hereafter the National Plan) [7], released in July 2021, describes a transition from ‘phase A’ in which strong suppression and no community transmission is the goal, to ‘phase B’ whereby SARS-CoV-2 infection was allowed to establish in the broader population. This transition was enabled by vaccination. The aim was for COVID-19 to be manageable, from both a public health and clinical perspective, through continued but more targeted application of public health and social measures (PHSMs) and test–trace–isolate–quarantine (TTIQ) strategies. The targets in the National Plan were agreed by all Australian States and Territories at National Cabinet (forum for the Prime Minister, state Premiers and Chief Ministers to meet) on 6 August 2021. The transition from phase A to phase B was the first-step on the path to ‘re-opening’. Latter phases ‘C’ and ‘D’ in the National Plan were supported by higher vaccine coverage levels allowing for further reductions in whole-of-population pandemic responses. Exploring target coverage thresholds for vaccination to enable the transition from phase A to phase B, and the required level of supporting interventions including ongoing PHSMs and TTIQ capabilities was identified as a priority under the National Plan. More broadly, identifying such thresholds was a common challenge for low incidence settings other than Australia [8, 9].

Here we report on the dynamic modelling study used to identify priority age groups for vaccination, target coverage thresholds, and the anticipated requirements for continued PHSMs and TTIQ to support a transition to a more open society in which SARS-CoV-2 could circulate while the health and clinical impacts remain manageable. Findings from this research directly informed specific re-opening policies within the National Plan and in particular the threshold and conditions for the transition from phase A to phase B.

## MODELLING FRAMEWORK

The modelling framework comprises three distinct components (depicted in Figure 1), each summarised here and described in detail in the Supplementary Material and references therein.

**Figure 1.**
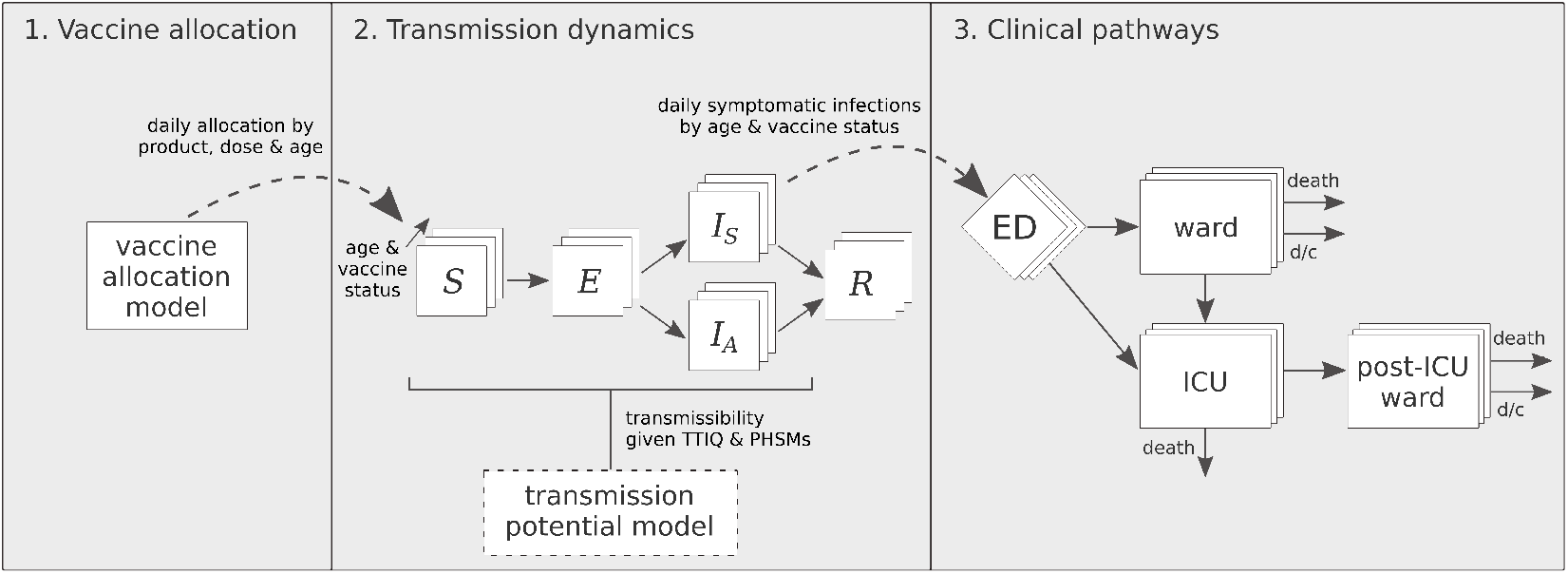
Schematic of three interlinked model components. 1. A model of vaccine allocation and roll-out to the Australian population (aged 16 and over). 2. An age-structured individual-based model of SARS-CoV-2 transmission dynamics to model infection numbers by age and vaccine-status. Note that transmissibility of SARS-CoV-2 under different PHSM bundles and TTIQ capabilities is informed by outputs from a model of SARS-CoV-2 transmission potential described in [10]. 3. A clinical-pathways stochastic model to map infection numbers to hospital admissions, ward and ICU occupancy, and deaths stratified by age and vaccine-status. *S* = susceptible; *E* = exposed; *I*_*S*_ = infectious and develops symptoms; *I*_*A*_ = infectious and does not develop symptoms; *R* = recovered; ED = emergency department, ICU = intensive care unit; d/c = discharge.

- A model of vaccine allocation and roll-out to the Australian population (aged 16 and over);
- An age-structured individual-based model of SARS-CoV-2 transmission dynamics to model infection numbers by age and vaccination-status; and
- A clinical-pathways stochastic model to map infection numbers to hospital admissions, ward and intensive care unit (ICU) occupancy, and deaths.

These three components are underpinned by an analysis of SARS-CoV-2 transmissibility in Australia, measured by the ‘transmission potential’ [11] (*TP*). As described elsewhere [12], the *TP* draws on a number of behavioural data streams to estimate (at a state level) the reproduction number that could be expected during widespread transmission. The *TP* is distinct from the effective reproduction number in that it represents the expected reproduction number of a pathogen in the general population rather than the reproduction number among active cases (who may not be representative of the general population). The *TP* is routinely reported for all states and territories of Australia and publicly available through the Australian Government’s Common Operating Picture [13]. Under pre-pandemic conditions, the *TP* corresponds to the basic reproduction number, *R*_0_. It varies through time due to changes in health system performance, different public health orders, and trends in population mixing and behaviours (such as limiting non-household contacts and adherence to cough etiquette and other infection control recommendations). Furthermore, because vaccines act to reduce transmission (through a reduction in the probability of acquisition and reduced contagiousness given breakthrough infection), additional reductions in *TP* due to vaccination can be estimated in the context of behavioural and public health response settings.

Using the *TP* model and time-series data on cases and population behaviours in Australia since March 2020, we first conducted a static analysis (described in [10]), to estimate *TP* for the Delta variant achieved under alternate vaccine allocation strategies, PHSMs, and TTIQ capabilities. An example output from this analysis is displayed in Figure 2 (adapted from [10]).

**Figure 2.**
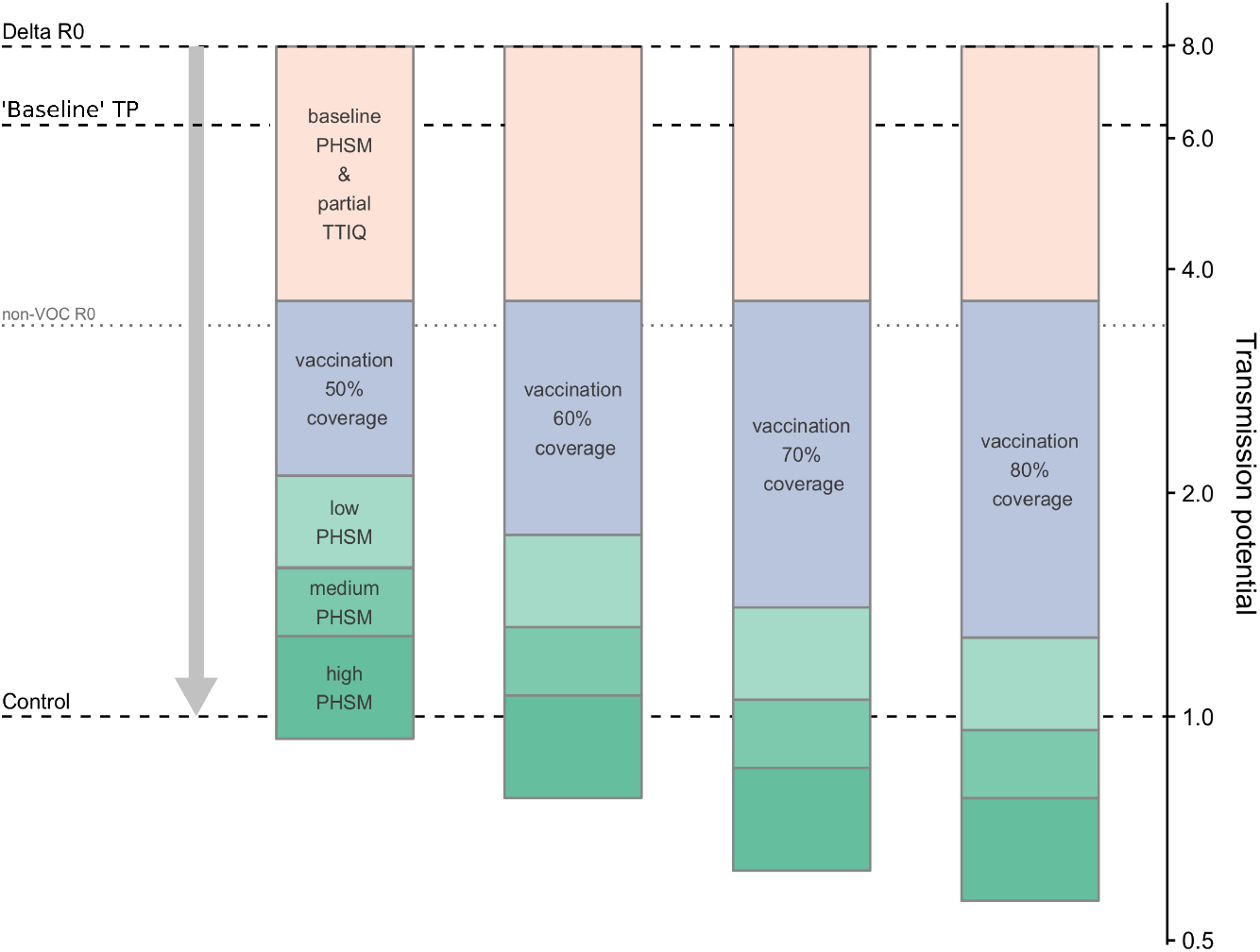
Figure adapted from [10] showing the combined effects of vaccination and PHSMs on ‘transmission potential’ (*TP*) of the SARS-CoV-2 Delta variant assuming partial TTIQ effectiveness and implementation of the ‘Transmission reducing’ vaccine allocation strategy. The dashed line,’Baseline’ *TP* indicates the baseline reproduction number of 6.32 for all scenarios considered in this study (prior to the application of TTIQ, enhanced PHSMs, and vaccination). Interventions are assumed to have multiplicative impact, and so a log scale is appropriate.

For the static analysis, we considered four ‘bundles’ of PHSMs: baseline, low, medium, and high (as shown in Figure 2). Each bundle relates to a specific time and place in Australia’s pandemic experience, thereby capturing both behavioural responses and the proportional reduction in *TP* achievable by PHSMs in the Australian context (see Table 1 for details).

**Table 1.** Description of measures implemented under different ‘bundles’ of public health and social measures (PHSMs). Each bundle relates to a specific time and place in Australia’s pandemic experience up to mid-2021 — thereby capturing behavioural responses and the proportional reduction in *TP* achievable by PHSMs in the Australian context. The proportional reductions in *TP* observed at each time and place can therefore be related to similar reductions achieved via other combinations of PHSMs (not limited to the “bundles” in place during the reference period). Similarly, the imposition of any given combination of PHSMs at different times and places may result in variable population responses and thus reductions in *TP*.

| PHSM bundle | Description |
| --- | --- |
| Baseline | Minimal density/capacity restrictions and no major outbreaks as in NSW March 2021 |
| Low | More stringent capacity restrictions compared to baseline (*e.g.*, hospitality venues limited to 10 customers per booking), as in NSW 23 August 2020 |
| Medium | Stringent capacity restrictions, group size limits (*e.g.*, fewer than five people outdoors), stay-at-home orders (except work, study, essential purposes), as in NSW 1 July 2021 |
| High | No household visitors, curfew, stay-at-home orders (except essential purposes and permitted work), schools closed (remote learning only), as in VIC 23 August 2020 |

The static analysis also considered the contribution of TTIQ in reducing virus spread, which is described in detail in [10]. Briefly, this included a detailed study of a limited time-series of case data from New South Wales between July 2020 and January 2021. During this time, case loads in New South Wales were low and the impact of TTIQ was clear in suppressing transmission [14, 12]. The empirical distribution of times from case detection to isolation was determined and used to evaluate the reduction in transmission due to TTIQ. This reduction — 54% — defines an ‘optimal’ TTIQ effect, representing the maximum reduction in *TP* we may expect due to TTIQ at any time during an epidemic. By assuming improvements in TTIQ are proportional to improvements in times to detection (*i.e*., times from symptom onset to test), the relative performance of TTIQ was measured during different periods of epidemic activity. This approach provided a distribution of times to isolation in epidemiological contexts where TTIQ was assessed to be partially effective. In particular, when calibrated against the data for Victoria from 4 August 2020 — the peak of daily locally-acquired COVID-19 cases in Australia in 2020 — this gives an estimated reduction of 43% in *TP* and defines the ‘partial’ TTIQ scenarios explored in the static analysis.

For all scenarios considered in this study, the dynamic transmission model uses a baseline reproduction number of 6.32, corresponding to a *TP* estimated for the Delta variant under baseline PHSM and after ‘backing out’ the effect of TTIQ (indicated by the dashed line ‘Baseline TP’ in Figure 2). The baseline *TP* is lower than the *R*_0_ for SARS-CoV-2 due to the baseline behavioural changes in the Australian population. It is further reduced based on assumed impact of (partial or optimal) TTIQ, enhanced (low, medium or high) PHSMs, and vaccination.

Here, we examine epidemic dynamics and clinical consequences of infections following transition from phase A to B of the National Plan (Table 2) at different vaccine coverage thresholds between 50% and 80%, for alternative age-based vaccine prioritisation strategies, and assuming continued application of PHSMs and TTIQ.

**Table 2.** Phases of the “National Plan to transition Australia’s National COVID-19 Response“[7]. Our modelling analysis focuses on the transition from ‘phase A’ in which strong suppression and no community transmission is the goal, to ‘phase B’ where vaccine coverage is high and SARS-CoV-2 infection is allowed to establish in the population. Scenarios therefore examine the epidemic dynamics and clinical consequences of infections following seeding of an epidemic at different vaccination prioritisation strategies and coverage thresholds.

| Phase | Description | Activities |
| --- | --- | --- |
| A | Vaccinate, Prepare and Pilot | Continue to strongly suppress the virus for the purpose of minimising community transmission |
| B | Vaccination Transition | Seek to minimise serious illness, hospitalisation and fatality as a result of COVID-19 |
| C | Vaccination Consolidation | As above |
| D | Post-Vaccination | Manage COVID-19 consistent with public health management of other infectious diseases |

### Vaccine allocation strategy and timing of the roll-out

Vaccine prioritisation in the Australian population through the first half of 2021 was based on a direct-protection approach, targeted towards those most at risk of severe outcomes. Two products were approved for distribution: AstraZeneca (ChAdOx1 nCoV-19) and Pfizer/BioNTech (BNT162b2 (mRNA)). By 11 July, based on Australian Immunisation Register (AIR) data, 33% of the population had received one-dose and 11% two-doses of a licensed vaccine (details provided in Table S1). This low starting point provided substantial scope to explore the importance of age cohort coverage within overall targets of 50-80% uptake in the population aged 16 years and over.

From this baseline position, four vaccine allocation strategies, and associated delivery scenarios, were considered: ‘Oldest first’, ‘40+ years first’, ‘All adults’, and an implementable strategy consistent with the national COVID-19 immunisation programme designated the ‘Transmission reducing’ strategy. Table 3 provides details for these alternative allocation strategies. The population-level impacts of these strategies are strongly related to underlying assumptions for age-specific mixing, susceptibility and infectiousness. A detailed description of these assumptions and their interaction with differential age cohort coverage under the target thresholds is provided in [10].

**Table 3.**
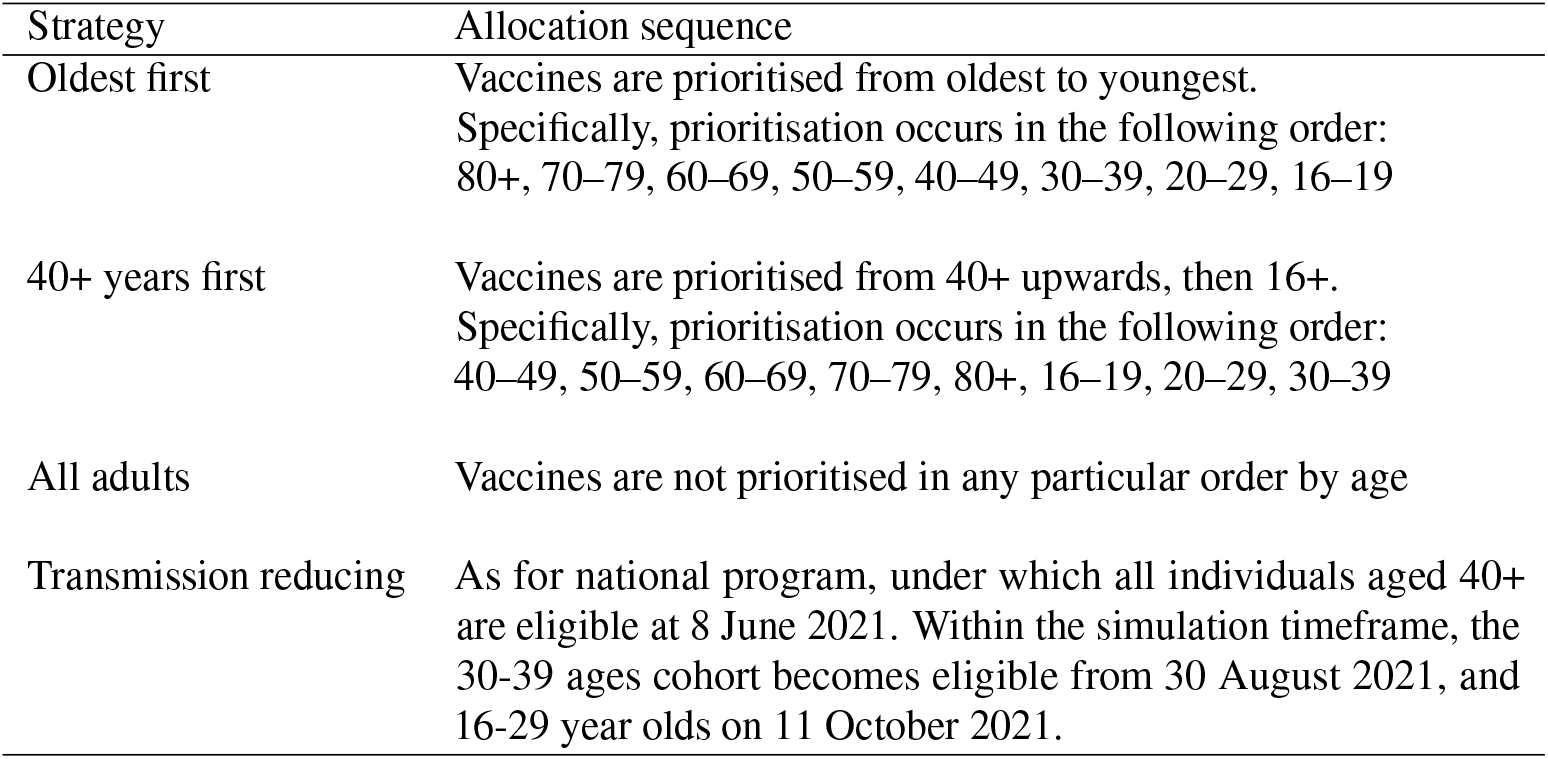
Vaccine allocation strategies by age, assuming July 2021 recommendations for AstraZeneca vaccine age eligibility (60+ years) and dosing interval (12 weeks)

Within the constraints of available supply (at the time of analysis, July/August 2021), AstraZeneca is provided to those aged 60+ years at a dosing interval of 12 weeks. Pfizer/BioNTech is provided to those aged 16–60 years at a dosing interval of three weeks. For both vaccines, a two week delay from second dose completion to full efficacy was assumed.

The rate of vaccine roll-out, and cumulative coverage by age-group, is modelled using an agent-based model utilising location and allocation data on vaccination sites and location data for the Australian population. In brief, each week a subset of the population seeks vaccination (according to the allocation strategy’s age-based eligibility criteria) at available sites within their respective geographic area. Sites receive deliveries of vaccines and administer vaccinations to the seeking population up to their level of stock. Figure S1 presents the modelled two-dose vaccine coverage time-series by age group under the four allocation strategies described above and Table S2 displays modelled terminal vaccine coverage by age group and strategy. In all scenarios considered below, the vaccine allocation model outputs full time-series by vaccine type (Pfizer, AstraZeneca) and dose (dose 1 and dose 2), which are fed into the transmission dynamics model.

For any given coverage threshold considered for the transition from phase A to phase B in the National Plan, the *TP* is computed (methods detailed in [10]). This differs by allocation strategy as vaccine product (Pfizer, AstraZeneca) varies by age, and the timing between doses varies for Pfizer and AstraZeneca. Table 4 presents the achieved *TP* at the key eligible-population (16+ years) coverage thresholds of 50%, 60%, 70% and 80%. These values define the initial transmissibility of SARS-CoV-2 in subsequent simulations of epidemic activity as now described. Vaccination continues to roll-out during these simulations according to the mean modelled output from the allocation model.

**Table 4.** Transmission potential for different vaccine allocation strategies and threshold coverage levels (16+ ages) for transition from phase A to phase B. These values assumed baseline PHSMs and partial TTIQ.

| Strategy | Coverage threshold |  |  |  |
| --- | --- | --- | --- | --- |
|  | 50% | 60% | 70% | 80% |
| Oldest first | 2.1 | 1.7 | 1.5 | 1.3 |
| 40+ years first | 2.1 | 1.9 | 1.6 | 1.3 |
| All adults | 2.0 | 1.7 | 1.5 | 1.3 |
| Transmission reducing | 2.1 | 1.8 | 1.4 | 1.3 |

### Transmission model

We developed an age-structured individual-based model (IBM) of SARS-CoV-2 transmission dynamics, calibrated to the Australian population. Panel 2 of Figure 1 presents a simplified state-diagram of the epidemiological-status for an individual in the synthetic population (see Supplementary Material for a more detailed schematic). Individuals in the population may be susceptible to infection, partially protected due to vaccination (with multiple sub-classes depending on vaccine-type, number of doses received and time since last vaccination), exposed, infectious or recovered. Among those infectious, the model distinguishes between those displaying symptoms or otherwise.

Age-specific mixing, susceptibility, and transmissibility assumptions employed in the dynamic transmission model are the same as those used in the static analysis [10]. Briefly, population mixing within and between age groups is configured based on synthetic social contact matrices published by [15], expanded to include an 80+ age class (assumed to have the same mixing rates as 75–79 years). Estimates of age-specific susceptibility and symptomatic fractions from [16] are used to compute an age-specific transmission matrix calibrated to the population-wide *TP* (Figure S3). Of note, the greatest transmission intensities are anticipated between individuals aged from 15–24 years, remaining high through adults of working age. While intense school-based mixing is anticipated between children aged 5–14, the transmission matrix accounts for the relatively low observed infectiousness of this age group, associated with a high proportion of asymptomatic infections. The age-specific contributions to *TP* accounting for demography, relative susceptibility and transmissibility and vaccine coverage for overall coverage levels (16+) of 50%, 60%, 70% and 80% under the four allocation strategies (oldest first, 40+ years first, all adults, and transmission reducing) are examined in [10].

COVID-19 vaccines have been shown to act on multiple elements of transmission and disease. Vaccine parameters related to transmission and symptomatic disease — for the Delta variant — are detailed in Tables 5 and 6, respectively. We assume that vaccination reduces susceptibility to infection (according to Table 5, left column) and the probability of developing symptomatic disease given infection (according to Table 6). The latter impacts transmission since we assume that asymptomatic individuals are 50% less infectious. We further assume that infected vaccinated individuals are less infectious by a factor calculated to match combined vaccine effectiveness assumptions on transmission (Table 5, right column). We use a baseline reproduction number of 6.32, corresponding to a *TP* estimated for the Delta variant under ‘baseline’ PHSMs and after ‘backing out’ the effect of TTIQ (indicated by the dashed line ‘Baseline *TP*’ in Figure 2).

**Table 5.** Vaccine parameters — transmission

| Vaccine | Reduction in susceptibility ($E_i$) | Reduction in contagiousness ($E_t$) | Overall reduction<sup>†</sup> |
| --- | --- | --- | --- |
| Astra Zeneca dose 1 | 18% | 48% | 57% |
| Astra Zeneca dose 2 | 60% | 65% | 86% |
| Pfizer dose 1 | 30% | 46% | 62% |
| Pfizer dose 2 | 79% | 65% | 93% |

<sup>†</sup> The overall reduction in transmission due to vaccination is given by $1 - (1 - E_i)(1 - E_t)$

**Table 6.** Vaccine parameters — clinical outcomes

| Vaccine | Vaccine effectiveness against |  |  |  |
| --- | --- | --- | --- | --- |
|  | Symptomatic infection<sup>a</sup> | Hospitalisation<sup>b</sup> | ICU admission<sup>c</sup> | Mortality<sup>b</sup> |
| Astra Zeneca dose 1 | 33% | 69% | 69% | 69% |
| Astra Zeneca dose 2 | 61% | 86% | 86% | 90% |
| Pfizer dose 1 | 33% | 71% | 71% | 71% |
| Pfizer dose 2 | 83% | 87% | 87% | 92% |

<sup>a</sup> Sheik et al [6]. Study reports VE against asymptomatic and symptomatic infection. We use their estimates of VE against symptomatic infection.
<sup>b</sup> London School of Hygiene and Tropical Medicine (LSHTM) central estimates used for UK roadmap modelling on 9 June 2021 for Delta, see Table 3 [4]. These Delta VE assumptions are scaled from VE estimates for pre-existing and Alpha variants. The starting Alpha assumptions for *hospitalisation* and *second dose mortality* are based on a range of studies and are in line with Public Health England's (PHE) COVID-19 vaccine surveillance report for pre-Alpha and Alpha (week 22) [19]. The starting Alpha assumptions for *first dose mortality* are informed by findings from Dagan et al [20] and Lopez Bernal et al [21]. Note that these assumptions are consistent with PHE's week 31 report (5 August 2021) [22]. To obtain estimates for Delta, the Alpha VE assumptions for both hospitalisation and mortality were reduced by half of the relative reductions by dose and product estimated by Lopez Bernal et al [23] for symptomatic infection (see Table 2). See LSHTM roadmap report from 9 June for further details [4].
<sup>c</sup> Few studies report VE against ICU admission for either ancestral or Delta variants. One study conducted in India (Victor et al [24]) reports 95% and 94% reductions in ICU admission after dose 1 and dose 2 of AstraZeneca, respectively. The findings from this study are unlikely to be directly transferable to the Australian setting due to health system differences. In the absence of relevant data for our setting, we assume the same reductions in ICU admission given vaccination as for hospitalisation.

The static analysis of vaccine impacts on *TP* [10] indicated that even 80% coverage under an optimal allocation strategy was unlikely to achieve a control *TP* of 1. We therefore consider the overlaid impacts of differing degrees of PHSMs and TTIQ in the dynamic transmission model. The application of low and medium PHSMs are considered in the IBM through modification to the baseline reproduction number as estimated by [10]. Partial and optimal TTIQ are incorporated by sampling the isolation time for each infected individual from the corresponding distribution of times from infection to isolation as estimated by [10].

We seed simulations with a fixed low number of infections at the time the vaccine coverage threshold is reached, reflecting a scenario in which the virus re-establishes itself in the Australian population via a border incursion or is allowed to enter given the achieved vaccine threshold. We seed with a sufficient number of infections (30 unvaccinated people by default) to exclude (with high probability) the chance of stochastic extinction.

Despite Australia’s strong suppression strategy, in late June 2021 the Delta variant had established itself in the community in New South Wales and Victoria. By late August 2021 (the time of these analyses) daily case incidence was in the thousands (New South Wales) and hundreds (Victoria), and were zero or low in other jurisdictions. Accordingly, we also examined scenarios in which the initial number of infections is ‘low’ (tens of infections), ‘medium’ (hundreds of infections) or ‘high’ (thousands of infections). While each of these initial conditions is low by global standards, they are highly pertinent to the Australian context and re-opening plan. For these medium and high initial conditions, we seed epidemics with a mix of vaccinated and unvaccinated individuals. Our approach to initialisation is described in detail in the Supplementary Material. Of note, cumulative incidence remains negligible in both cases at the time of reaching the coverage threshold and so scenarios remain equivalent in terms of the susceptible population size. To aid comparisons in this additional analysis of initial seeding size, we also re-run the ‘low’ scenario, simulating tens of infections in both vaccinated and unvaccinated individuals.

Further technical details of the IBM construction are provided in the Supplementary Material.

### Clinical pathways

Similar to previous studies [17, 18], we model hospital admissions, ward/ICU occupancy and death. The model takes inputs of daily symptomatic infections, stratified by age and vaccine status, from the transmission dynamics model. A fraction of those with symptomatic infection, either vaccinated or unvaccinated, will present to hospital and require admission for additional care. Patients admitted to hospital may occupy either a ward or ICU bed. Ward stays may also deteriorate and require ICU care, before returning to a general ward and discharge. Death as an endpoint may also occur while admitted, during either a ward or ICU stay. These flows are represented using a stochastic model, with age-specific transitions and length-of-stay distributions informed by international data. Details are provided in the Supplementary Material. Vaccination parameters related to clinical outcomes were calibrated to the Alpha variant due to lack of available data on Delta at the time this research was conducted. We note that around the time of initial reporting of our findings to policymakers, evidence of Delta’s increased severity (in both vaccinated and unvaccinated individuals) was beginning to emerge, and we reflect on the consequences of this in the Discussion. Table 6 provides the vaccine parameters related to clinical outcomes. Of note, our analysis does not account for health system capacity constraints, and so outputs represents the anticipated demand for clinical services. Thus if a scenario were to exceed health system capacity, our simulations would underestimate clinical burden since individuals who are unable to access care will likely have worse outcomes.

### Graphical presentation

Figures displaying time-series of epidemiological quantities computed from the transmission and clinical pathways models show the 90% confidence interval of trajectories calculated across time (*i.e*., the 5- and 95-percentiles on each day) as coloured ribbons. A single, representative, trajectory is also shown to give an indication of the dynamics within each scenario. The single trajectory within each scenario was selected as corresponding to the median total infections (or symptomatic infections, ward occupancy, ICU occupancy, deaths, as appropriate) across the projection horizon.

## RESULTS

A comparison of time-series for daily infections, by vaccine allocation strategy and vaccine coverage threshold for transition from phase A to B, is presented in Figure 3. The corresponding time-series for ward and ICU occupancy, and deaths, are displayed in Figures S6–S9, and cumulative infections and deaths are displayed in Figures S10 and S11. Here, baseline PHSMs and partial TTIQ are assumed to be in place. These analyses demonstrate that the ‘All adults’ and ‘Transmission reducing’ strategies result in fewer infections (Figures 3 and S10) and afford greater clinical protection (Figures S7, S8, S9, and S11) compared to the ‘Oldest first’ and ‘40+ years first’ strategies at all transition coverage levels. Furthermore, marked reductions in the time to epidemic peak and peak size are noted as vaccine coverage increases from 50 to 60% and beyond.

**Figure 3.**
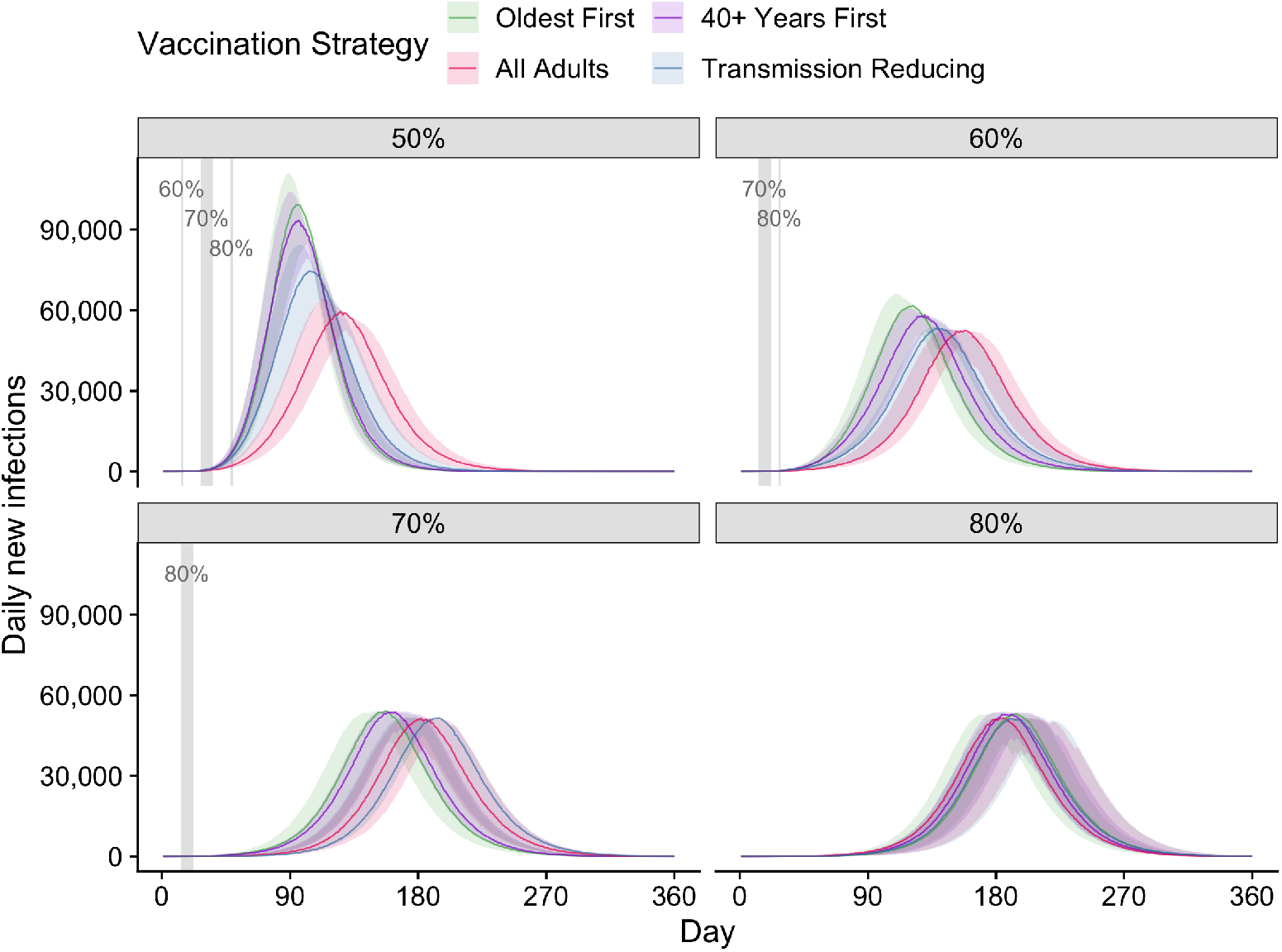
Comparison of time-series for infections (asymptomatic and symptomatic) by vaccine allocation strategy and vaccine coverage threshold for transition from the National Plan phase A to B. All simulations were seeded with a fixed, low number of infections (30 unvaccinated people). Solid coloured lines and coloured shading represent median epidemic trajectories and 95% confidence intervals respectively. Vaccination continues to roll-out beyond the target threshold during each simulation according to the mean modelled output from the allocation model. Solid grey vertical lines (or shading) indicate the date of achieving each vaccine coverage threshold.

The ‘Transmission reducing’ strategy was designed as an implementable version of the ‘All adults’ strategy and is considered from herein. In recognition of the Delta variant becoming established in some Australian jurisdictions prior to reaching 70% coverage thresholds, we now examine the consequences of an increased case load at the time of transition from phase A to phase B in the National Plan. We examine the role that overlaid PHSMs and/or more effective TTIQ may play in reducing epidemic activity, particularly given the context of likely higher initial case incidence.

Row one of Figure 4 presents time-series of infections (asymptomatic and symptomatic combined) for three initial conditions following transition from phase A to phase B at either 70% (left) or 80% (right) coverage. Baseline PHSMs and partial TTIQ are applied. For a transition at 70% coverage, an increase from ‘low’ (tens) to ‘medium’ (hundreds) numbers of infections at the time of transition results in a marked leftward shift in the timing of the epidemic. This result is unsurprising given basic epidemic theory. However, if the transition occurs at a point with ‘high’ (thousands) infections, the epidemic is not only left-shifted, but peak and final size also increase. This is a result of dynamic ‘overshoot’, otherwise avoidable due to the continued roll-out of vaccines and hence increasing coverage level during the epidemic. If the transition from phase A to phase B is made at 80% coverage, the leftward shift in epidemic dynamics remains, but even for the ‘high’ (thousands) infections scenario, there is minimal impact on peak and overall size of the epidemic. This is due to two factors: 1) vaccine coverage is sufficiently high to prevent a large ‘overshoot’, and 2) under the model for vaccine distribution, the rate of increase in coverage slows beyond 80% as we approach saturation of vaccine uptake in the eligible population (around 89.9% of the eligible population, reached at 56 weeks). Corresponding time-series for clinical outcomes are presented in rows two to four of Figure 4.

**Figure 4.**
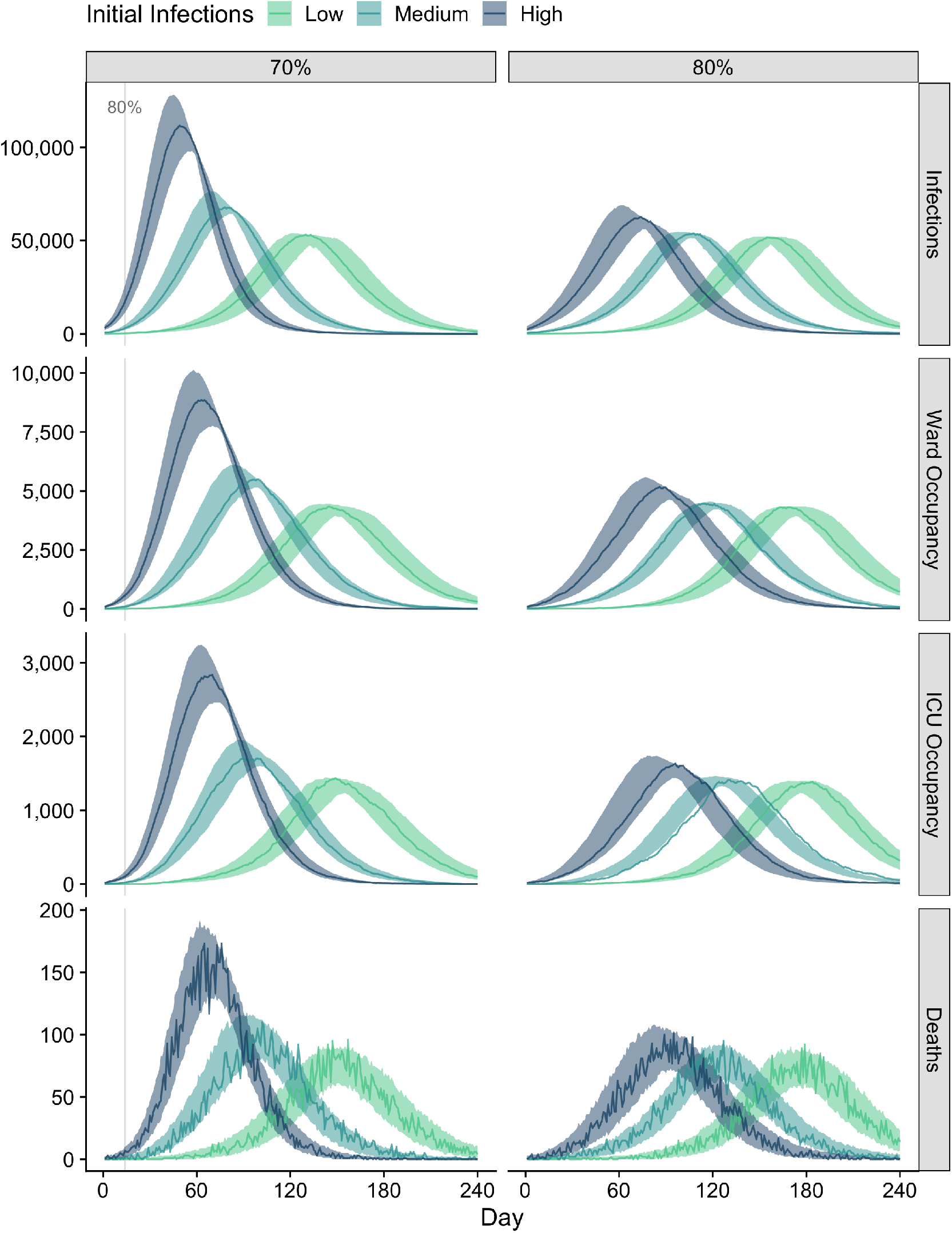
Time-series of infections (asymptomatic and symptomatic), occupied ward and ICU beds, and deaths for epidemics seeded with low (tens), medium (hundreds), and high (thousands) numbers of initial infections at the 70% (left) and 80% (right) coverage thresholds. All scenarios assume baseline PHSMs and partial TTIQ are applied. Solid coloured lines and coloured shading represent median epidemic trajectories and 95% confidence intervals respectively. Vaccination continues to roll-out beyond the target threshold during each simulation according to the mean modelled output from the allocation model. Solid grey vertical lines indicate the date of achieving the 80% vaccine coverage threshold.

Australia’s National Plan envisages continued application of enhanced PHSMs to further suppress epidemic activity and minimise morbidity and mortality. Panels A and B of Figure 5 demonstrate the marked benefit of continued application of low PHSMs, accompanied by partial TTIQ, from the point of transition from phase A to phase B. At both 70% (panel A) and 80% (panel B) the epidemic is strongly suppressed compared to under baseline PHSMs (Figure 4). Similar beneficial outcomes can be achieved through application of optimal TTIQ under baseline PHSMs (Figure S12), although we note that maintaining optimal TTIQ was not considered feasible over the long term.

**Figure 5.**
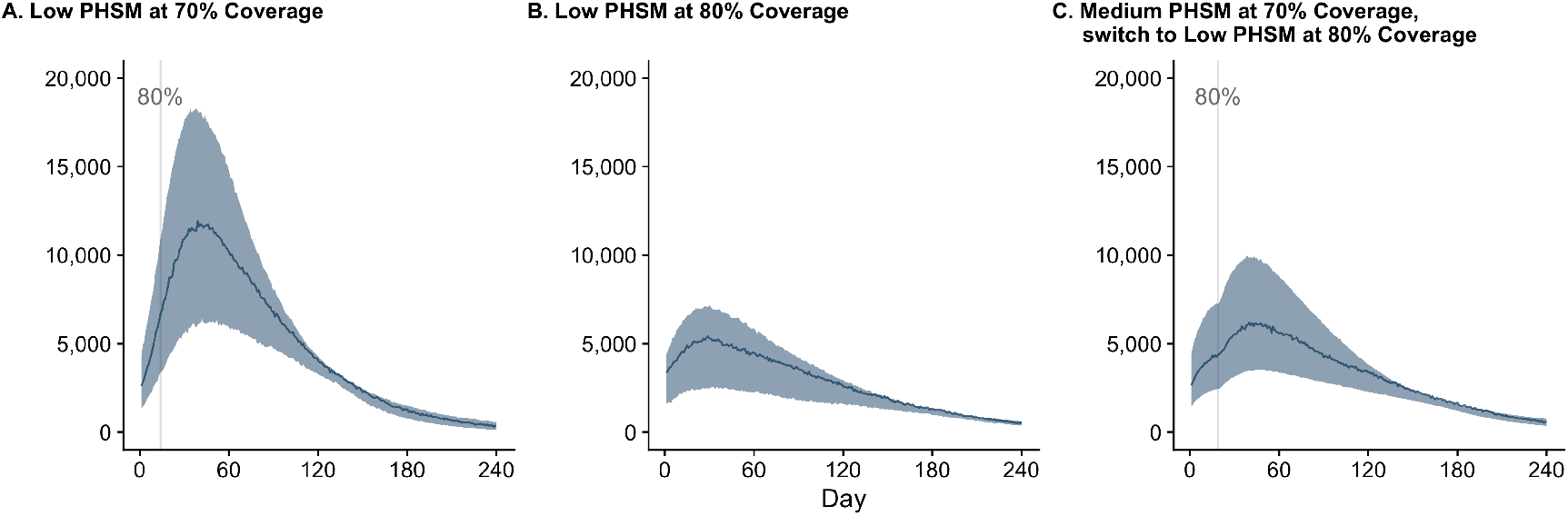
Time-series of infections (asymptomatic and symptomatic) when low PHSMs are applied from the point of the transition from phase A to B of the National Plan at 70% and 80% vaccine coverage (panels A and B). Panel C demonstrates an adaptive strategy where medium PHSMs are applied during the period from 70% to 80% coverage, easing to low PHSMs thereafter. All scenarios assume ‘high’ numbers of initial infections and the application of partial TTIQ. Solid coloured lines and coloured shading represent median epidemic trajectories and 95% confidence intervals respectively. Vaccination continues to roll-out beyond the target threshold during each simulation according to the mean modelled output from the allocation model. Solid grey vertical lines indicate the date of achieving the 80% vaccine coverage threshold.

While all of the above simulations assume a fixed policy from the point of transition from phase A to phase B, significant benefits may be realised by the imposition of increased restrictions for limited periods of time. Panel C of Figure 5 shows how an adaptive strategy can support a transition at 70% even under ‘high’ numbers of infections. Medium PHSMs are applied during the period from 70% to 80% coverage, with low PHSMs enacted thereafter. Compared to the scenario in which low PHSMs are active during this transition period (panel A), infections are notably reduced. Overall epidemic impact is similar to when the transition from phase A to phase B is only made at 80% (panel B).

Figure 6 compares the cumulative number of symptomatic infections and clinical impacts by age group and vaccine status for outbreaks seeded at 50% and 80%, assuming baseline PHSMs and partial TTIQ. There is a significant overall reduced burden given establishment of community transmission at 80% compared to 50% coverage (panels A and B). In this context, a substantial fraction of symptomatic infections and severe outcomes are anticipated in vaccinated individuals within highly vaccinated age groups. At both 50% and 80% vaccine coverage, a decoupling of infections from clinical burden is evident, with infections concentrated in younger less vaccinated populations, yet much greater health impacts are anticipated in older more highly vaccinated age groups (panels C and D).

**Figure 6.**
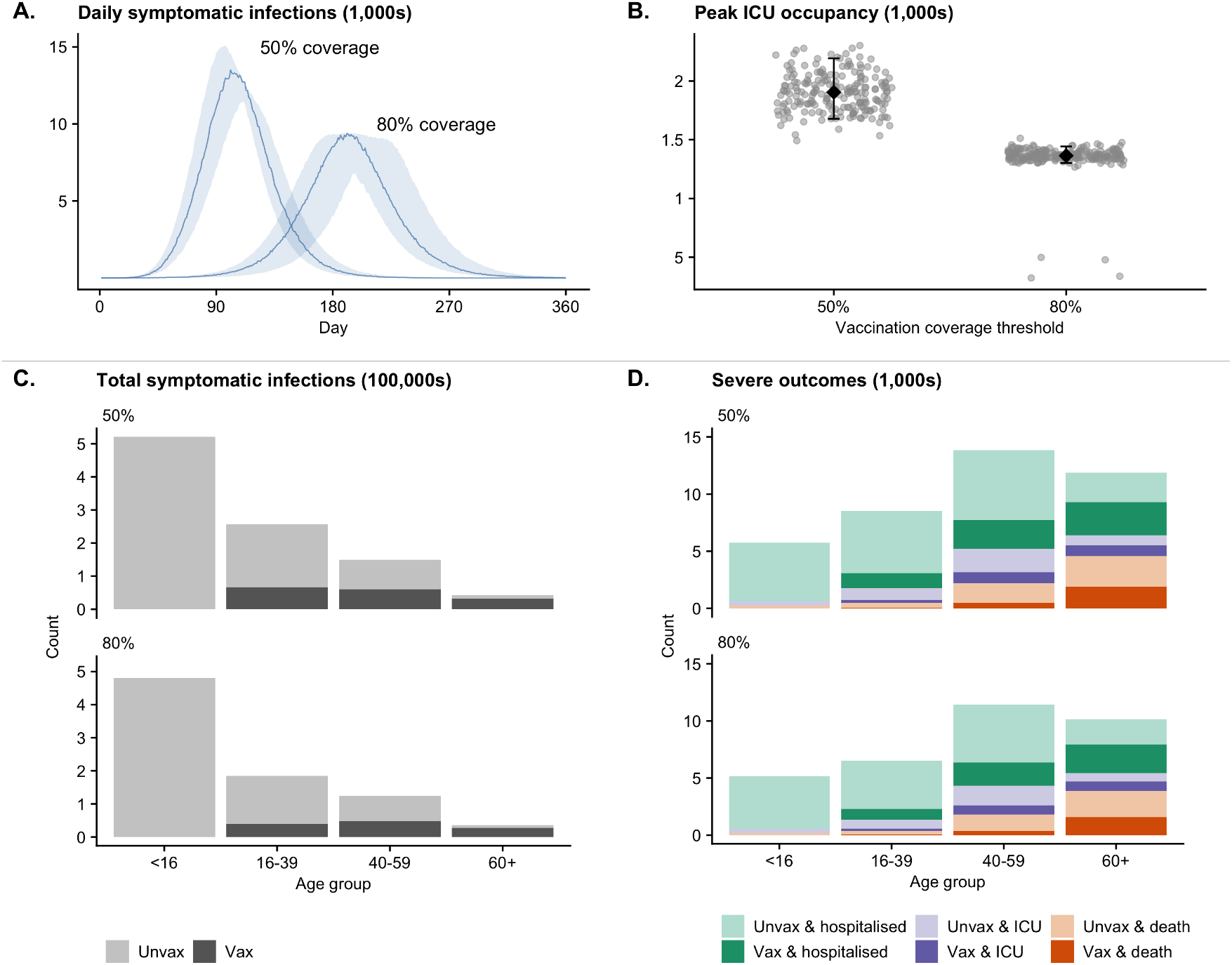
Comparison of symptomatic infections and clinical impacts given transition from phase A to B leading to established community transmission at coverage thresholds of 50% and 80% for the ‘Transmission reducing’ strategy, assuming baseline PHSMs and partial TTIQ. All simulations were seeded with a fixed, low number of infections (30 unvaccinated people). Vaccination continues to roll-out beyond the target threshold during each simulation according to the mean modelled output from the allocation model. Panel A: time-series of symptomatic infections (solid lines = median epidemic trajectory; shading = 95% confidence intervals). Panel B: Peak ICU occupancy (grey dots = estimate for each model simulation, black diamonds and bars = median and 5^th^ and 95^th^ percentiles). Panel C: Number of cumulative symptomatic infections by age group and vaccine status. Panel D: Number of cumulative ward admissions, ICU admissions and deaths by age group and vaccine status. Unvax = unvaccinated. Vax = vaccinated.

## DISCUSSION

To transition from a strategic goal of ‘no community transmission’ to one of ‘minimising COVID-19 burden’ requires sufficient vaccine coverage to 1) suppress case incidence such that TTIQ remains an effective response to reduce transmission; and 2) keep anticipated clinical loads within capacity. Here, through a model-based analysis, we have demonstrated that for Australia adult vaccine coverage needed to be at least 70% to support that transition. Our analyses were conducted based on assumed circulation of the Delta variant of SARS-CoV-2 and contingent upon the Australian epidemiological, health system and societal context in which circulation of SARS-CoV-2 was strongly suppressed up to the point of transition.

If a transition away from the goal of ’no community transmission’ were to occur when coverage was below 70%, our model predicts that virus transmission would escalate rapidly, either overwhelming clinical systems or requiring re-imposition of strong and broad ‘lockdown’ style responses (and even then with commensurate and avoidable infections and morbidity). If, as was the case in the state of Victoria, case incidence was high (thousands per day) at the time of reaching the 70% coverage threshold, then heightened but temporary public health and social measures (medium PHSMs) could help bridge the period to achieving 80% coverage, reducing the risk of a surge in transmission that may threaten capacity. With coverage at 80% or beyond, our model indicates that epidemic dynamics would likely be manageable within the constraints of the clinical system and be compatible with Australia’s strategic road-map for managing COVID-19 in the vaccine era.

Substantial gains — in terms of reduced cases and markedly reduced hospitalisations and deaths — could be afforded by maintenance of low PHSMs over the vaccine roll-out phase and beyond. Furthermore, those low PHSMs would support a strong and more effective TTIQ response, helping avoid escalation of local epidemic activity. If the transition to ‘living with COVID’ were to occur prior to reaching 70% coverage, case numbers would likely rise to such a level that TTIQ effectiveness was diminished and epidemic ‘overshoot’ would result in additional—and *a priori* avoidable—cases, hospitalisations, and deaths.

Our findings are comparable to those from model-based studies for other low prevalence settings [8, 9]. Nguyen *et al*. [8] and Steyn *et al*. [9] investigated the impacts of alternative age-based vaccine allocation strategies and coverage thresholds in the New Zealand context. Similar to Australia, New Zealand’s vaccine roll out was intended to support a shift in response strategy from elimination to border re-opening and virus circulation. Both studies concluded that high vaccine uptake (*e.g*., greater than 80% of the population aged 16+) and maintenance of other public health measures during the vaccine roll out phase would be required to prevent serious adverse health impacts. Furthermore, studies from high prevalence settings, including the United Kingdom, also highlighted the potential adverse health and health system impacts of complete relaxation of social restrictions during the early phases of vaccine roll out [4].

A key limitation of our work is that we considered a single, large population (24 million) in which the virus spreads. This was a deliberate and necessary choice designed to support the Australian national (whole-of-country) re-opening plan. However, and particularly in the early establishment phase of a country wide epidemic, transmission was expected to be highly focal. Jurisdictions where SARS-CoV-2 transmission established prior to the 70% coverage threshold, such as New South Wales and Victoria, began a transition from a state of ‘medium’ and ‘high’ case incidence respectively (Figures S13 and S14). Other jurisdictions maintained zero case incidence well beyond vaccine coverage thresholds of 70 and 80%. Furthermore, at a sub-jurisdictional level, we would expect systematic differences in vaccine coverage, behavioural patterns and TTIQ capabilities. These considerations emphasised the need for small-area assessment of *TP* (to anticipate risk) and other real time epidemiological metrics.

More broadly, it is not the goal of scenario analyses (such as those conducted here) to precisely predict the future course of the epidemic, nor impacts of specific policies, not least because of uncertainty in key model inputs, such as vaccine effectiveness, intrinsic severity of the Delta variant, and future population behaviour. When we reported the findings documented in this manuscript to government in mid-2021 to support Australia’s COVID response strategy, ongoing situational assessment (as described elsewhere [12, 14]) was acknowledged as critical to the success of the National Plan. That is, monitoring of local data was anticipated to allow bench-marking of the scenarios to guide real-time policy decision making on the transition to Phase B of the National Plan. Likewise, a summary of the scenario modelling on vaccination and the easing of restrictions in the United Kingdom, published in February 2021, articulated the need for measures to be relaxed “based on data and the situation at the time, rather than at pre-determined dates” [4].

The degree of PHSMs needed for disease control prior to the vaccine threshold being reached in Australia and during the transition period would require reference to near-real-time estimates of the effective reproduction number, and forecasts of cases and clinical burden, at a sub-national level. Outcomes were anticipated to be highly situation specific — related to the actual starting number of cases, the population characteristics where transmission is concentrated (*e.g*., vaccine coverage, age, co-morbidities, access to health services, ability to adhere to personal protective measures *etc*.), the rate of vaccination, and the level of epidemic suppression achieved.

In mid-2021 transmission of the Delta variant became established prior to the 70% coverage threshold in the jurisdictions of New South Wales, Victoria, and the Australian Capital Territory [25]. Informed by the model-based analyses presented here, state governments imposed strict stay-at-home measures (corresponding to high PHSMs) before relaxing those measures (to settings corresponding to approximately medium and then low PHSMs) upon reaching the 70 and 80% coverage thresholds respectively. Assisted by the ongoing application of PHSMs and TTIQ, and greater than 80% vaccine coverage, the initial waves of Delta infection had peaked and were in decline by November 2021, with epidemic activity stabilizing at levels manageable within health system capacity — as anticipated by the scenarios explored here.

Response plans, and the modelling work supporting them, should be adaptable to new phases of the pandemic, including the emergence of new variants and intervention options. All of the findings from our scenario analysis were made in the context of the Delta variant (specifically, what was known in June 2021) and the Australian healthcare system and society under conditions of low prevalence. In the latter half of 2021, the modelling framework described here was adapted to investigate new epidemic dynamics and policy needs in response to emerging information on the increased severity of Delta (relative to ancestral strains), the emergence of Omicron, and the roll out of third vaccine doses.

The Omicron variant (BA.1) was first detected in Australia in late November 2021 and rapidly became the dominant circulating variant. At the time, daily case incidence in five of Australia’s eight states/territories was either zero or fewer than tens of cases per day (Figures S13 and S14). Consequently, when widespread transmission became established at various time points beyond the 70% coverage threshold in late 2021/early 2022, the first ever SARS-CoV-2 epidemics managed by these jurisdictions were dominated by Omicron. Additional scenario analyses required adjustment to consider emerging evidence of Omicron’s increased intrinsic transmissibility, higher propensity for immune-evasion, and decreased clinical severity relative to the Delta variant. Furthermore, our vaccine allocation scenarios were restricted to two-dose vaccination of the 16+ adult population. With approval of vaccines for those 5–16 years of age and third doses for the adult population by late 2021, additional research was required to assess the anticipated additional benefits of these programs.

The scenario modelling described here informed a step-wise and agile approach to the relaxation of COVID-19 response measures in Australia in the vaccine-era.

## Data Availability

No new datasets are presented in this research. Code to perform the analyses is available at

https://github.com/aus-covid-modelling/NationalCabinetModelling

## DATA AVAILABILITY

No new datasets are presented in this research. Code to perform the analyses is available at: https://github.com/aus-covid-modelling/NationalCabinetModelling

## ACKNOWLEDGEMENTS

This work was directly funded by the Australian Government Department of Health Office of Health Protection. Additional support was provided by the Australian Research Council (NG DECRA fellowship DE180100635) and the National Health and Medical Research Council of Australia through its Centres of Research Excellence (SPECTRUM, GNT1170960) and Investigator Grant Schemes (JMcV Principal Research Fellowship, GNT1117140; FMS Emerging Leader Fellowship, 2021/GNT2010051). Computations were supported by: use of the Nectar Research Cloud, a collaborative Australian research platform supported by the National Collaborative Research Infrastructure Strategy (NCRIS); MASSIVE HPC facility (www.massive.org.au); and The University of Melbourne’s Research Computing Services.

## SUPPLEMENTARY MATERIAL

### Vaccine allocation strategy and timing of the roll-out

**Figure S1.**
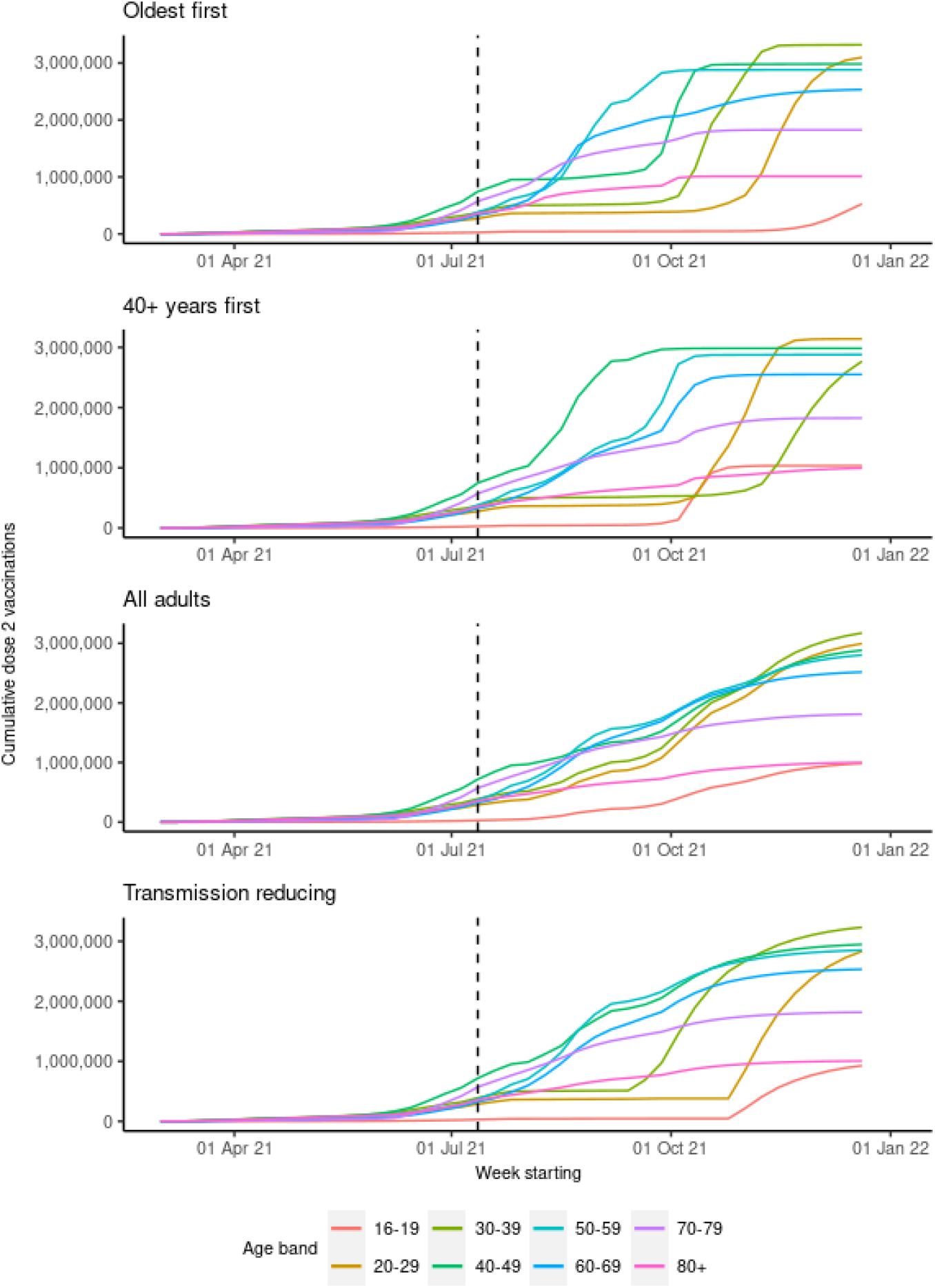
Time-series for modelled two-dose vaccination coverage by age group for the four strategies ‘Oldest first’, ‘40+ years first’, ‘All ages’, and ‘Transmission reducing’. Vertical dotted line indicates the starting point of modelled vaccinations.

**Table S1.** Distribution of vaccine coverage within each age band up to and including 11 July 2021 based on Australian Immunisation Register (AIR) data as of 15 July 2021.

| Age band | Pfizer |  | Astra Zeneca |  | Total |  |
| --- | --- | --- | --- | --- | --- | --- |
|  | Dose 1 | Dose 2 | Dose 1 | Dose 2 | Dose 1 | Dose 2 |
| 16–19 | 2.9% | 1.6% | 0.9% | 0.5% | 3.8% | 2.1% |
| 20–29 | 8.5% | 6.0% | 2.1% | 1.5% | 10.6% | 7.5% |
| 30–39 | 11.4% | 8.1% | 2.6% | 1.8% | 13.9% | 9.9% |
| 40–49 | 26.4% | 20.2% | 3.5% | 2.4% | 29.9% | 22.6% |
| 50–59 | 12.2% | 7.3% | 29.5% | 5.0% | 41.8% | 12.3% |
| 60–69 | 5.2% | 4.2% | 53.0% | 8.0% | 58.2% | 12.2% |
| 70–79 | 4.5% | 3.0% | 72.5% | 27.2% | 77.0% | 30.2% |
| 80+ | 16.2% | 11.8% | 65.3% | 22.0% | 81.5% | 33.8% |
| Total | 11.4% | 6.7% | 21.8% | 4.8% | 33.2% | 11.4% |

**Table S2.**
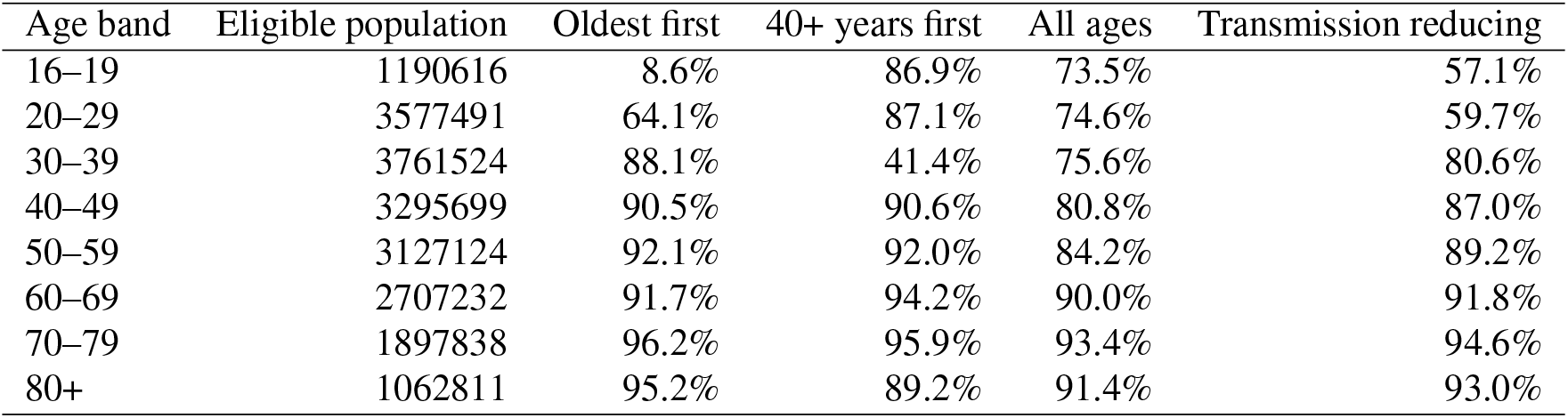
Distribution of vaccine coverage by age band by achievement of the 80% vaccine coverage threshold (1 November 2021) for standard Astra Zeneca dosing indications (60+, 12 week interval between doses) and the four vaccine allocation strategies.

| Age band | Eligible population | Oldest first | 40+ years first | All ages | Transmission reducing |
| --- | --- | --- | --- | --- | --- |
| 16–19 | 1190616 | 8.6% | 86.9% | 73.5% | 57.1% |
| 20–29 | 3577491 | 64.1% | 87.1% | 74.6% | 59.7% |
| 30–39 | 3761524 | 88.1% | 41.4% | 75.6% | 80.6% |
| 40–49 | 3295699 | 90.5% | 90.6% | 80.8% | 87.0% |
| 50–59 | 3127124 | 92.1% | 92.0% | 84.2% | 89.2% |
| 60–69 | 2707232 | 91.7% | 94.2% | 90.0% | 91.8% |
| 70–79 | 1897838 | 96.2% | 95.9% | 93.4% | 94.6% |
| 80+ | 1062811 | 95.2% | 89.2% | 91.4% | 93.0% |

### Transmission model

The purpose of the individual based model (IBM) developed in this work is to model the spread of COVID-19 through the Australian population, accounting for the effects of vaccination, PHSMs and TTIQ on transmission. Further, by combining the symptomatic infection trajectories from the IBM with a clinical pathways model, we can assess the anticipated burden of severe infections on the Australian health care system.

#### Entities and state variables

The main entity of interest in our model is the individual, which is used to represent a single resident within our population of interest. We use each individual’s internal state variables to govern the spread of COVID-19. The state variables assigned to our individuals are their age, age bracket, vaccine and disease status, time that they are isolated post infection and the number of secondary infections that they generate (see Table S3 for a summary of each state variable and entities). The vaccine status, time isolated and number of secondary infections are “empty” until the appropriate event triggers their initialisation.

Within each individual, the disease status and vaccine status entities are made up of their own internal state variables. The state variables of the disease status entity are the individual’s infection status, symptom status, probability of onward transmission, and time of exposure, as well as three timers for the individual’s time of symptom onset, time of infectious onset and time of recovery. The state variables of the vaccine status entity consists of the vaccine type and time of the most recent vaccination. Furthermore, we also store the probability of asymptomatic infection, susceptibility and probability of onward transmission corresponding to the most recent vaccine received by the individual.

**Table S3.** Description of state variables and entities of interest in the IBM.

| Entity | State variables | Description |
| --- | --- | --- |
| Individual | Age | The age of the individual in years. |
|  | Age bracket | Integer reference to the age bracket in the contact matrix. |
|  | Vaccine status | Entity containing the vaccine status of the individual. |
|  | Disease status | Entity containing information on the individual's disease state. |
|  | Time isolated | The time post infection that the individual will isolate. |
|  | Secondary infections | The number of people infected by this individual. |
| Vaccine status | Vaccine type | The type of vaccine this individual has recieved (AZ, Pfizer). |
|  | Dose number | The number of vaccine doses that the individual has received. |
|  | Time vaccinated | Time of most recent vaccination. |
| Disease status | Infection status | The current infection state of the individual (S, E, I, R). |
|  | Symptom status | Boolean for asymptomatic or symptomatic infection. |
|  | Onward transmission | Rate of onward transmission. |
|  | Time of exposure | The time the individual was exposed to COVID-19. |
|  | Time infectious | The time the individual becomes infectious. |
|  | Time of symptoms | The time that the individual will develop symptoms. |
|  | Time of recovery | The time that the individual will recover. |

#### Process overview and scheduling

The IBM consists of two main processes, namely, the vaccination schedule and the infection model. These processes are interrelated as vaccination leads to a change in the properties of each individual which is then propagated into the infection model, altering the spread of COVID-19. Both models are simulated simultaneously using a discrete time-step, *δt* days, with *t* = 0 coinciding with the start (15 February 2021) of the vaccination schedule provided by QUANTIUM (Figure S1).

The vaccination schedule within the IBM follows a predetermined roll-out as shown in Figure S1. This roll-out is coupled into the IBM using the following process. Within each time-step, we look-up the number of individuals in each age bracket (these age brackets differ from those used in the contact matrix) that will receive their first dose according to the vaccination roll-out. We then uniformly sample individuals from within the age brackets to vaccinate and update the state variable controlling their vaccine status. At this point in time, we also assign the time that these individuals will receive their second dose of the same vaccine product (this is vaccine product dependent).

**Figure S2.**
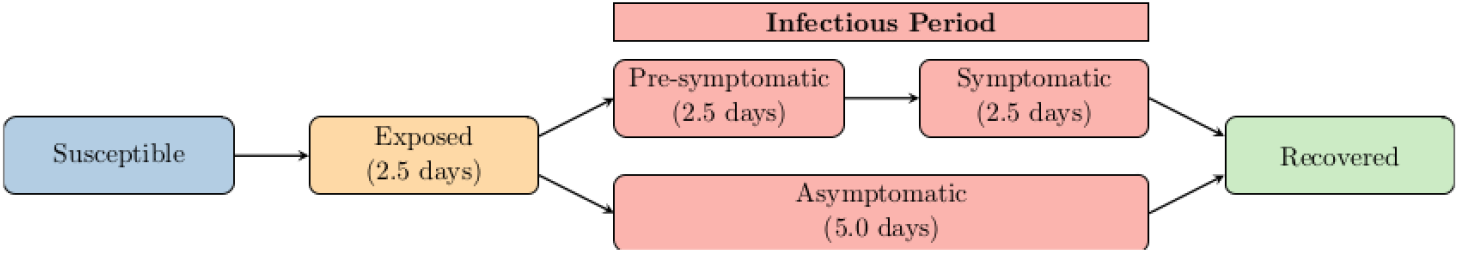
Transmission model: a simple schematic describing the mutually-exclusive states that an individual may reside in, and the transition between those states.

The infection model is based off a Susceptible-Exposed-Infected-Recovered (SEIR) paradigm. A simplified diagram of which is provided in Figure S2. In simple terms, the individuals in our simulation are susceptible until they are infected by an infectious contact, at which point they will transition into the exposed state. At a later time, the exposed individual will move into the infectious state and be pre-symptomatic. This is the period where the individual is infectious but is currently not displaying any symptoms. After a finite amount of time in pre-symptomatic state, the individual will become either asymptomatic or symptomatic. Finally, the individual will transition into the recovered state and no longer be infectious. Note that for our purposes the actual dynamics within each compartment are more complicated, depending upon all state variables of the individuals, the specifics of which will be detailed here.

To simulate infection within the IBM, we explicitly track the exposed and infected individuals in two dynamic arrays. It is only important to track the exposed and infected individuals as those that are susceptible and recovered do not directly contribute to the spread of COVID-19. At each time-step we update all individuals in the exposed dynamic array, followed by updating all individuals in the infected dynamic array, tracking all transitions between compartments for the next time-step.

For each of the individuals in the exposed dynamic array we determine whether or not they will become infectious at this point in time. This is done by checking against the member variable that contains the time they become infectious. If the individual becomes infectious at this time-step, they are removed from the exposed dynamic array and added to the end of the infected array for the next time-step (care must be taken to ensure that the newly infectious individual is not updated twice in a single time-step now that they are in the infected array).

The individuals in the infected dynamic array are updated as follows. We assume that the number of contacts made by each infectious individual is Poisson distributed, unless the internal state variable, time isolated, indicates that the infected individual is now successfully isolated. If the individual is isolated, we assume that they make no contacts. If the individual is not isolated, than the number of contacts, *C*_*i,k*_, made by an individual in age group *i* with individuals in age group *k* is,

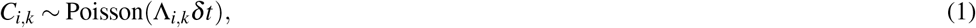

where Λ_*i,k*_ is the (*i, k*)th element of the contact matrix. Once we determine the number of contacts made in each age bracket, we sample who the infected individual will contact uniformly over the known population. For each of the susceptible contacts, we then determine if the contact resulted in a successful infection event.

The probability of infection given contact with a susceptible individual is

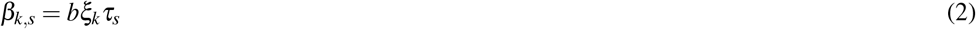

where *τ*_*s*_ is the transmissibility of the infector (depends upon symptom status *s*), *b* is the baseline probability of infection given contact and *ξ*_*k*_ is the relative susceptibility of the infectee (depending upon what age group they are in). To account for the effect of vaccination, 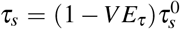 and 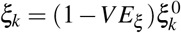 where 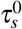 is 1 or 0.5 for symptomatic and asymptomatic individuals respectively, 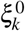 isthe baseline susceptibility for an individual in age bracket *k, VE*_*τ*_ is the vaccine efficacy against onward transmission and *VE*_*ξ*_ is the vaccine efficacy against infection. If the infection of the contact is successful, we increment the secondary infections state variable of the infected individual by one and expose the contact.

The exposure of a contact sets the appropriate state variables of the newly infected individual. To do so, we set their infection status to exposed and add their information to the dynamic array of exposed individuals. At this point, we set the individuals time of exposure and sample the time that the individual will transition to the infectious state, the time that they will develop symptoms, the time of isolation given symptom onset and the time that they will recover. Using the known vaccine status of the newly exposed individual, we determine if they will develop symptoms and assign their transmissibility for the entire infectious period. To determine if the individual is asymptomatic or symptomatic we use a Bernoulli trial with probability 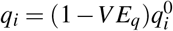, where *VE*_*q*_ is the vaccine efficacy against symptoms and 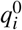 is thebaseline probability of symptoms for an individual in age bracket *i*. The transmissibility of the newly infected individual is defined at exposure and is assumed to remain constant throughout the individual’s entire infectious period.

Once all contacts and infection events have been simulated for the infectious individual, we check if they either become symptomatic or recover. To determine if an individual becomes symptomatic, we check if the symptom timer should be triggered. To determine if the individual recovers we check if the recovery timer has triggered. At the time of recovery, the infectious individual’s infection status is changed to recovered and they are removed from the dynamic array that tracks infectious individuals. It is assumed that once recovered, the individual will remain recovered for the rest of the simulation (no waning immunity). Note that due to the assumption of no waning immunity, we do not have to be vigilant in resetting any state variables within the individual. As they are recovered, we ensure that all further infection events in which they are involved fail. Finally, we store all statistics of the infection event for output at the end of the simulation.

#### Initialisation

To initialise the model, we create the population using data from the vaccine model, with each individual’s initial disease status being set to the susceptible state. We then calculate *b* in Equation 2 using the transmission potential for the spread of COVID-19, see the section ***Transmission potential in the individual-based model*** for a derivation of *TP* in the IBM.

For the initial scenarios shown in Figures 3, we seed 30 infections in the unvaccinated individuals on the day that the target coverage is achieved. In the scenarios with low, medium and high seeding at the target coverage (shown in Figures 6 and 5, we seed 30 infections in the unvaccinated individuals at a date in advance of achieving the target coverage. We then ensure that only simulations within the bounds of low (30-100 daily infections), medium (300-1000 daily infections) and high (1000-4500 daily infections) infections are considered.

#### Transmission potential in the individual-based model

We assume that the transmission potential (*TP*), as described in Ryan et al, is the expected number of secondary infections arising from the initial seeding of infection:

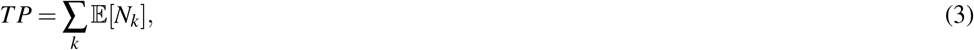

where *N*_*k*_ is the number of secondary infections in age group *k*. We split secondary infections based upon the symptom status and age group of the infected individual:

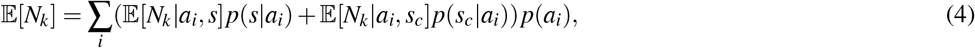

where *a*_*i*_ is the *i*th age group and *s* indicates symptoms and *s*_*c*_ indicates asymptomatic. The quantity *p*(*s*|*a*_*i*_) is the probability of symptoms given the individual is in age group *i*, while *p*(*s*_*c*_|*a*_*i*_) = 1− *p*(*s*| *a*_*i*_)| is the probability an individual of age *i* is asymptomatic. Finally, *N*_*k*_ *a*_*i*_, *s* is the number of secondary| infections for an individual in age group *i* with symptoms and *N*_*k*_ *a*_*i*_, *s*_*c*_ is the corresponding value for asymptomatic individuals.

Next, we determine expressions for 𝔼[*N*_*k*_ |*a*_*i*_, *s*]*p*(*s*| *a*_*i*_) and 𝔼[*N*_*k*_|*a*_*i*_,*s*_*c*_]*p*(*s*_*c*_| *a*_*i*_)). Eq. (1) defines the number of contacts in age bracket *k* made by an infectious individual in age bracket *i* in a single time step.Therefore, the total number of contacts made throughout an infectious period is distributed according to,

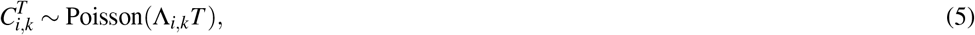

where 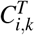 is used to the total number of contacts and *T* is the time the individual is infectious.

Using Eq. (2), we know that the total number of secondary infections in age bracket *k* caused by an infected individual in age bracket *k* follows a Binomial distribution, such that,

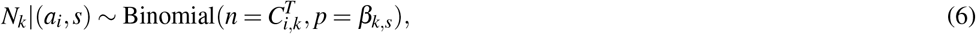

with an expected value of

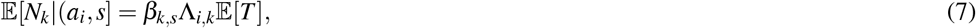

which has been obtained using the law of total expectation.

By combining Eq. (4) and Eq. (7) we obtain the expected number of secondary infections caused by an infectious individual in age bracket *i*, namely,

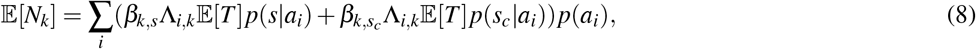

and we estimate the overall *TP* as

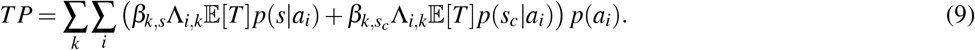

In our case, *β*_*k,s*_ = *bτ*_*s*_*ξ*_*k*_ with known *τ* and *ξ*. Therefore,

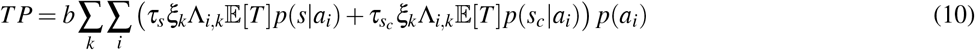

which is

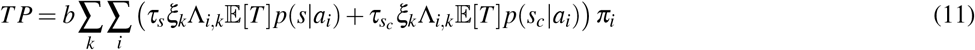

where *π*_*i*_ is the proportion of infected population in age bracket *i*. Rewriting *p*(*s*|*a*_*i*_) = *q*_*i*_ and *p*(*s*_*c*_|*a*_*i*_) = 1 − *q*_*i*_, where *q*_*i*_ is the proportion of symptomatic infections age group *i*, we get:

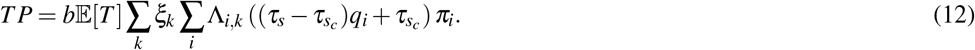

It is possible to solve for *b* given a supplied *TP*, and it is how we can match this model to a transmission scenario with a specified transmission potential.

**Figure S3.**
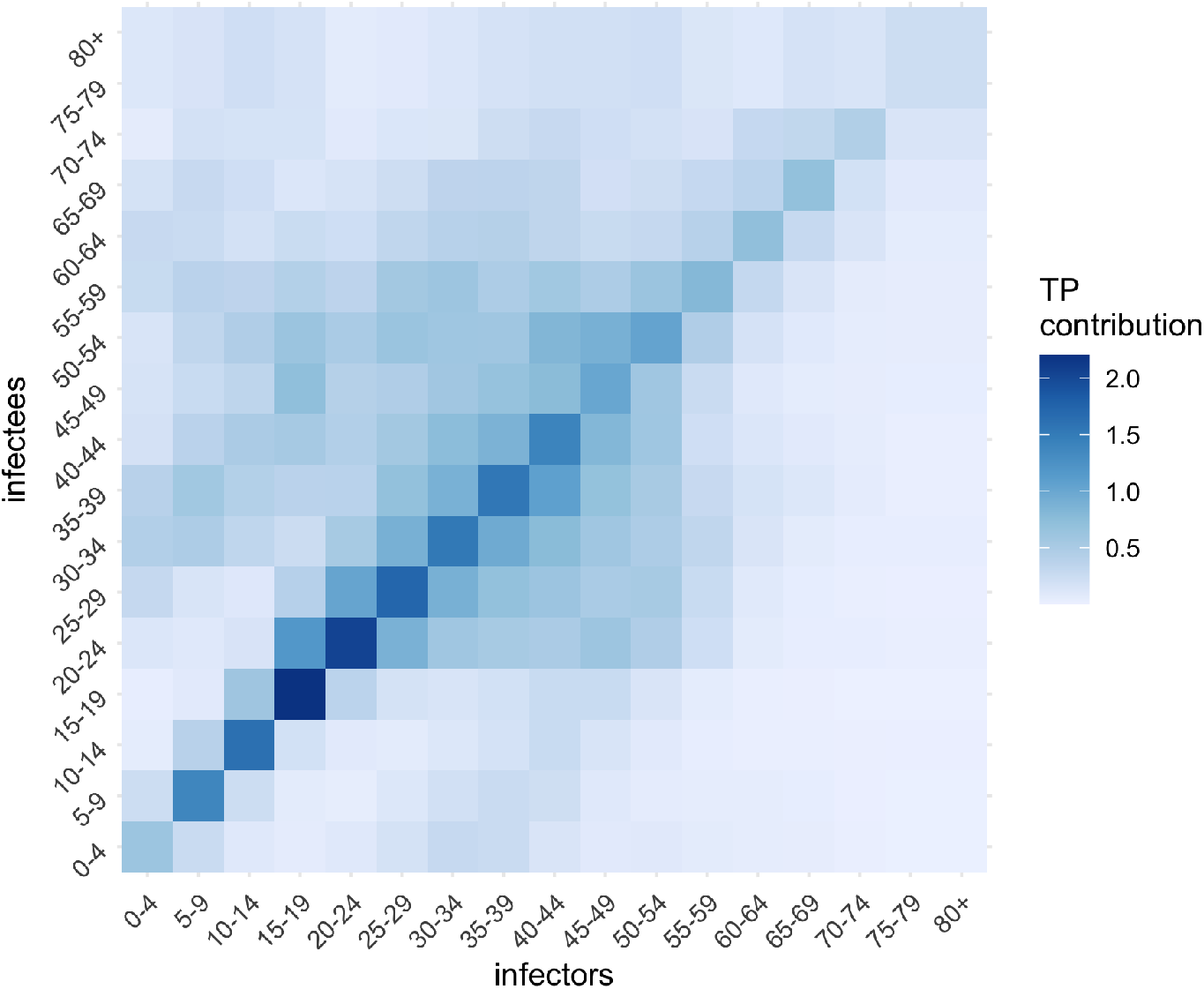
Age-based transmission matrix (assuming no vaccination) accounting for age-dependent mixing (derived from Prem et al. [15]) and age-dependent susceptibility and contagiousness.

### Clinical pathways

#### Overview

The clinical pathways model takes as input the daily number of symptomatic cases, stratified by vaccine status (dose number and product) and age, from the transmission model and outputs projected ward occupancy, ICU occupancy, and deaths over time. A simplified representation of this model is shown in Figure S4. There is a delay between the onset of symptoms and presenting at ED. Upon arrival at ED, the individual is assumed to be admitted to ward or ICU immediately. Individuals who are admitted to ward will either die, be discharged from ward, or require ICU care (this may occur on the same day as admission to ward). Individuals in ICU will either die in ICU or return to ward, from here they will either die or be discharged.

**Figure S4.**
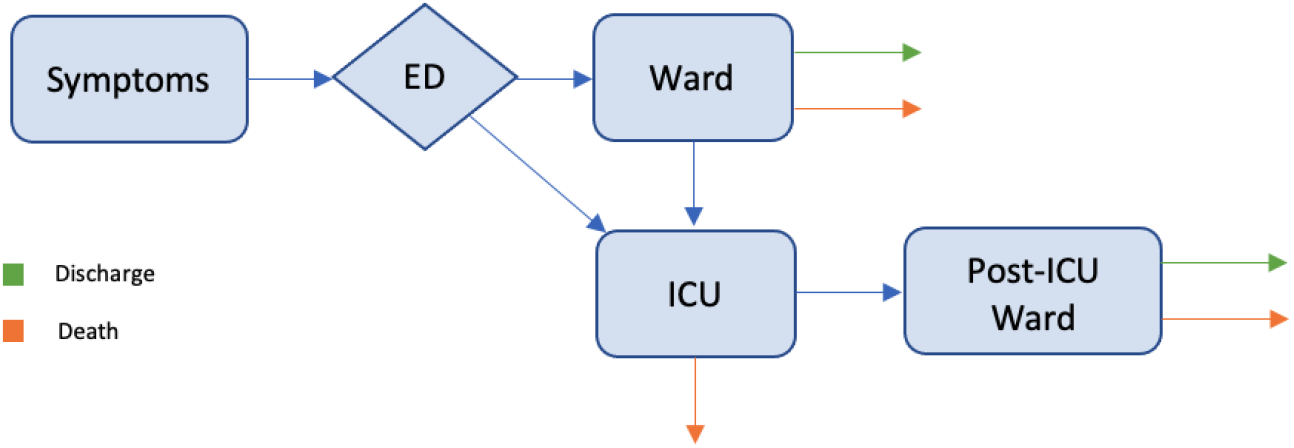
A simplified diagram of the clinical pathways model. ED = emergency department. ICU = intensive care unit.

The lengths of stay (LoS) in each compartment/clinical setting depends on the eventual clinical pathway of individuals. For example, lengths of stay in ward will typically be shorter for individuals who later require ICU care. The pathways of individuals through the health system are dependent on both their age and vaccination status. The LoS distributions are taken from an analysis of UK data from March–May 2020 [17], and therefore based on patients infected with ancestral strains of SARS-CoV-2. The age stratified transition probabilities between model compartments are taken from an analysis of SARS-CoV-2 infection severity by the same group [17], and then scaled for the Delta variant according to Table S6 and vaccination status according to Table 6.

### Model structure

The compartments in the clinical pathways model in their entirety are shown in Figure S5. Here, the ward, ICU, and post-ICU compartments are split into categories based on the eventual trajectories of individuals. By splitting compartments in this way, the model can account for differences in the LoS depending on the eventual outcomes of individuals. Blue, orange and green arrows in Figure S5 represent internal model transitions, deaths, and discharges respectively. Transition probabilities are dependent on the age and vaccination status of the individual (the probabilities of severe outcomes are higher for older and unvaccinated individuals). The model structure is broadly consistent with that described in [17] except that individuals are stratified by vaccination status and transition probabilities are scaled for the Delta variant and vaccination status.

Notation for the model compartments and transition probabilities are given in Tables S4 and S5. Further, the LoS distributions and the prior on the mean LoS are given in Table S4. Here, LoS for each compartment is assumed to be Gamma distributed with mean and shape parameters (which do not need to be integer). These LoS distributions are discretised over days. Events can happen in zero days, in particular, it is possible for hospital admission and entry to ICU to occur on the same day. The mean LoS for each simulation is sampled from a Gaussian distribution with the same mean and comparable variance reported in [17]. For the mean LoS in *H*_*R*_, *H*_*D*_ and *ICU*_*pre*_, the prior variance is taken to be zero as these estimates have negligible uncertainty in [17]. The time from symptom onset to hospital presentation uses the same distribution as [17] — an exponential distribution with mean 4 days.

**Figure S5.**
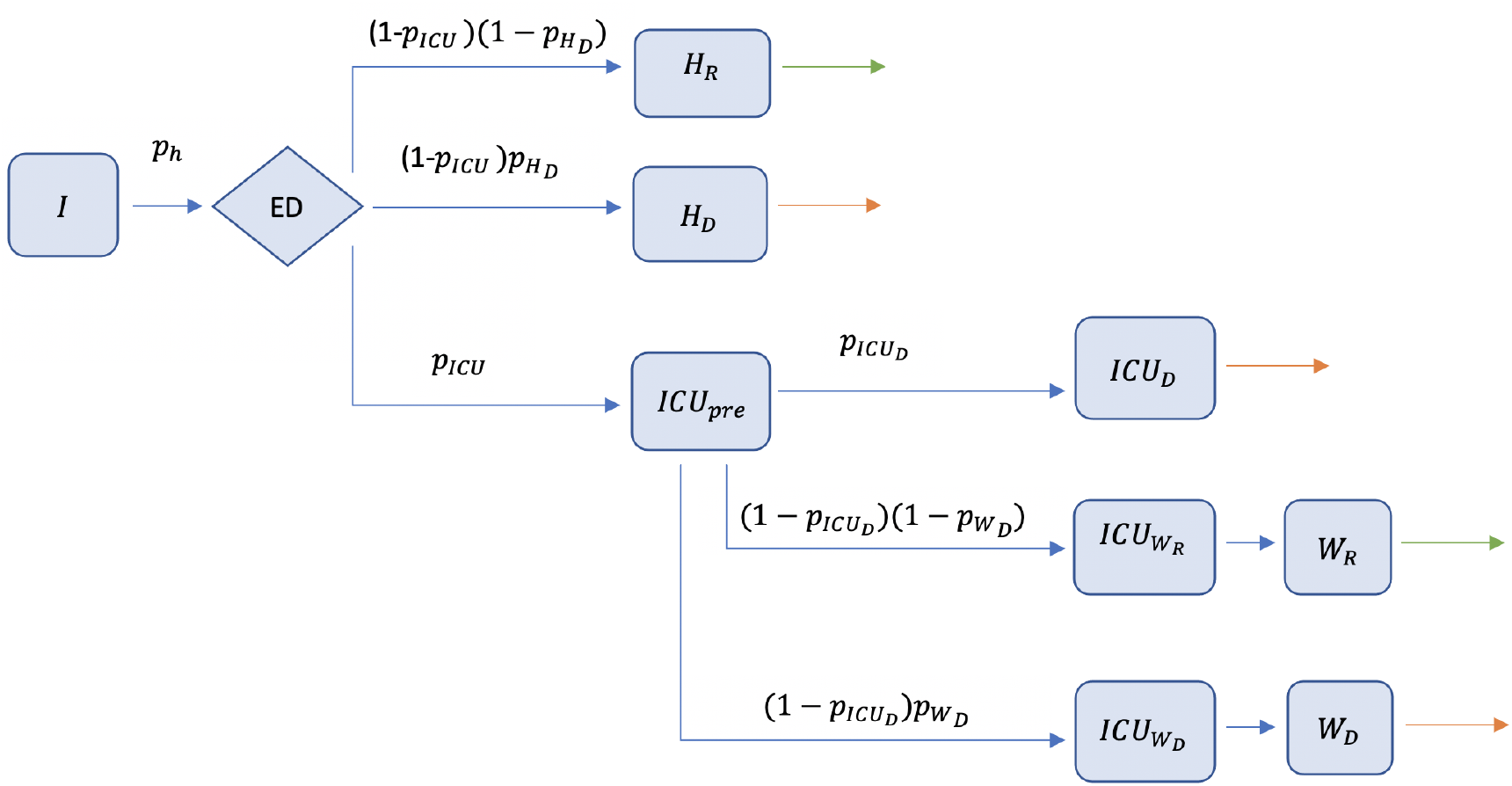
A diagram of all compartments in the clinical pathways model. Model compartment and transition probability descriptions are provided in Tables S4 and S5 respectively.

Table S7 gives the transition probabilities by age for unvaccinated individuals. These values are taken from [17] and scaled for the Delta variant according to Table S6. Note, the scaling of some severity parameters are given in terms of an odds ratio. That is, for baseline probability, *p*, and odds ratio, *r*, the adjusted probability, *p*^∗^, is given by

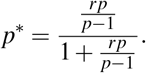

Similar scaling of the severity parameters for vaccination status are based on odds ratios presented in Table 6.

**Table S4.** Table of clinical pathways model compartments, their description, the parameters of their Gamma distributed length of time in each compartment, and the prior standard deviation on the mean length of stay (LoS).

| Compartment | Description | Mean LoS | Shape | Prior std dev. |
| --- | --- | --- | --- | --- |
| $H_R$ | Ward (non-ICU) that will recover | 10.7 | 1 | 0 |
| $H_D$ | Ward (non-ICU) that will die | 10.3 | 2 | 0 |
| $ICU_{pre}$ | Ward that will go to ICU | 2.5 | 1 | 0 |
| $ICU_D$ | ICU that will die in ICU | 11.8 | 2 | 0.10 |
| $ICU_{W_R}$ | ICU that will move to ward and recover | 15.6 | 1 | 0.26 |
| $ICU_{W_D}$ | ICU that will move to ward and die | 7 | 1 | 0.51 |
| $W_R$ | post-ICU ward that will recover | 12.2 | 2 | 0.10 |
| $W_D$ | post-ICU ward that will die | 8.1 | 1 | 0.97 |

**Table S5.** Table of clinical pathways model transition probabilities.

| Probability | Description |
| --- | --- |
| $p_h$ | Present at hospital given symptomatic infection |
| $p_{ICU}$ | ICU given admitted to ward |
| $p_{H_D}$ | Death on ward given admitted and will not go to ICU |
| $p_{ICU_D}$ | Death in ICU given admitted to ICU |
| $p_{W_D}$ | Death on post-ICU ward given admitted to post-ICU ward |

**Table S6.**
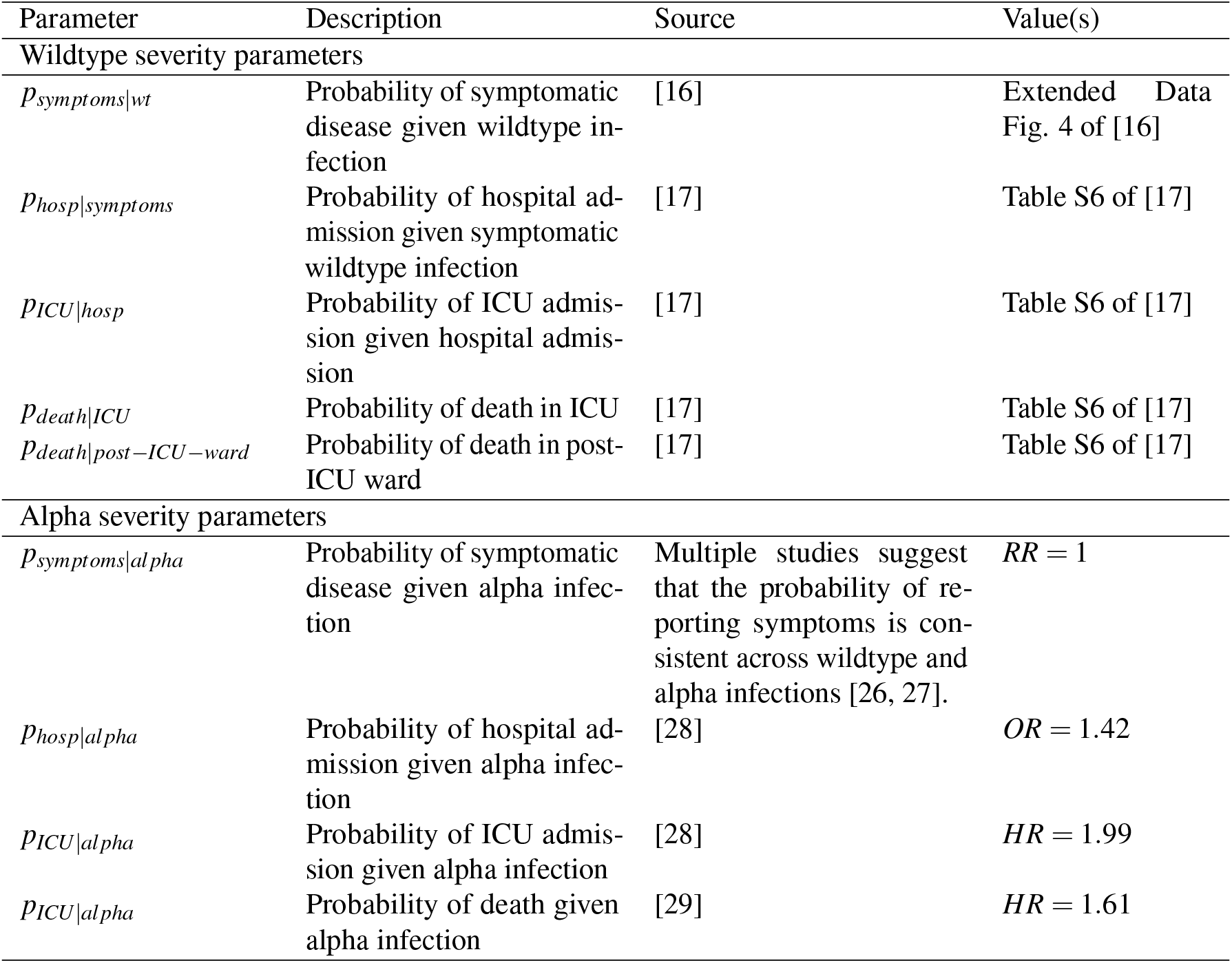
Disease severity assumptions for wildtype and the Alpha variant.

| Parameter | Description | Source | Value(s) |
| --- | --- | --- | --- |
| Wildtype severity parameters |  |  |  |
| $p_{symptoms\vert wt}$ | Probability of symptomatic disease given wildtype infection | [16] | Extended Data Fig. 4 of [16] |
| $p_{hosp\vert symptoms}$ | Probability of hospital admission given symptomatic wildtype infection | [17] | Table S6 of [17] |
| $p_{ICU\vert hosp}$ | Probability of ICU admission given hospital admission | [17] | Table S6 of [17] |
| $p_{death\vert ICU}$ | Probability of death in ICU | [17] | Table S6 of [17] |
| $p_{death\vert post-ICU-ward}$ | Probability of death in post-ICU ward | [17] | Table S6 of [17] |
| Alpha severity parameters |  |  |  |
| $p_{symptoms\vert alpha}$ | Probability of symptomatic disease given alpha infection | Multiple studies suggest that the probability of reporting symptoms is consistent across wildtype and alpha infections [26, 27]. | $RR = 1$ |
| $p_{hosp\vert alpha}$ | Probability of hospital admission given alpha infection | [28] | $OR = 1.42$ |
| $p_{ICU\vert alpha}$ | Probability of ICU admission given alpha infection | [28] | $HR = 1.99$ |
| $p_{ICU\vert alpha}$ | Probability of death given alpha infection | [29] | $HR = 1.61$ |

**Table S7.** Transition probabilities by age for unvaccinated individuals scaled for the Delta variant.

| | $p_h$ | $p_{ICU}$ | $p_{H_D}$ | $p_{ICU_D}$ | $p_{W_D}$ |
| --- | --- | --- | --- | --- | --- |
| [0, 5) | 0.041 | 0.082 | 0.021 | 0.152 | 0.024 |
| [5, 10) | 0.001 | 0.097 | 0.019 | 0.155 | 0.022 |
| [10, 15) | 0.006 | 0.113 | 0.018 | 0.157 | 0.020 |
| [15, 20) | 0.009 | 0.131 | 0.019 | 0.162 | 0.019 |
| [20, 25) | 0.027 | 0.150 | 0.019 | 0.168 | 0.019 |
| [25, 30) | 0.042 | 0.171 | 0.021 | 0.177 | 0.020 |
| [30, 35) | 0.044 | 0.194 | 0.025 | 0.191 | 0.021 |
| [35, 40) | 0.047 | 0.222 | 0.031 | 0.211 | 0.022 |
| [40, 45) | 0.052 | 0.258 | 0.043 | 0.241 | 0.024 |
| [45, 50) | 0.077 | 0.298 | 0.064 | 0.281 | 0.026 |
| [50, 55) | 0.140 | 0.334 | 0.099 | 0.327 | 0.029 |
| [55, 60) | 0.198 | 0.357 | 0.155 | 0.382 | 0.035 |
| [60, 65) | 0.244 | 0.352 | 0.234 | 0.436 | 0.048 |
| [65, 70) | 0.390 | 0.324 | 0.348 | 0.487 | 0.072 |
| [70, 75) | 0.565 | 0.260 | 0.483 | 0.525 | 0.110 |
| [75, 80) | 0.809 | 0.177 | 0.627 | 0.542 | 0.153 |
| 80+ | 0.729 | 0.045 | 0.679 | 0.497 | 0.216 |

### Supplementary results figures

Figures S6–S9 below show daily symptomatic infections, ward occupancy, ICU occupancy, and daily deaths, respectively, for each vaccination coverage threshold (50–80%), and each of the four vaccine allocation strategies (oldest first, 40+ years first, all ages, transmission reducing). Figures S10 and S11 show cumulative infections (asymptomatic or symptomatic) and deaths respectively.

**Figure S6.**
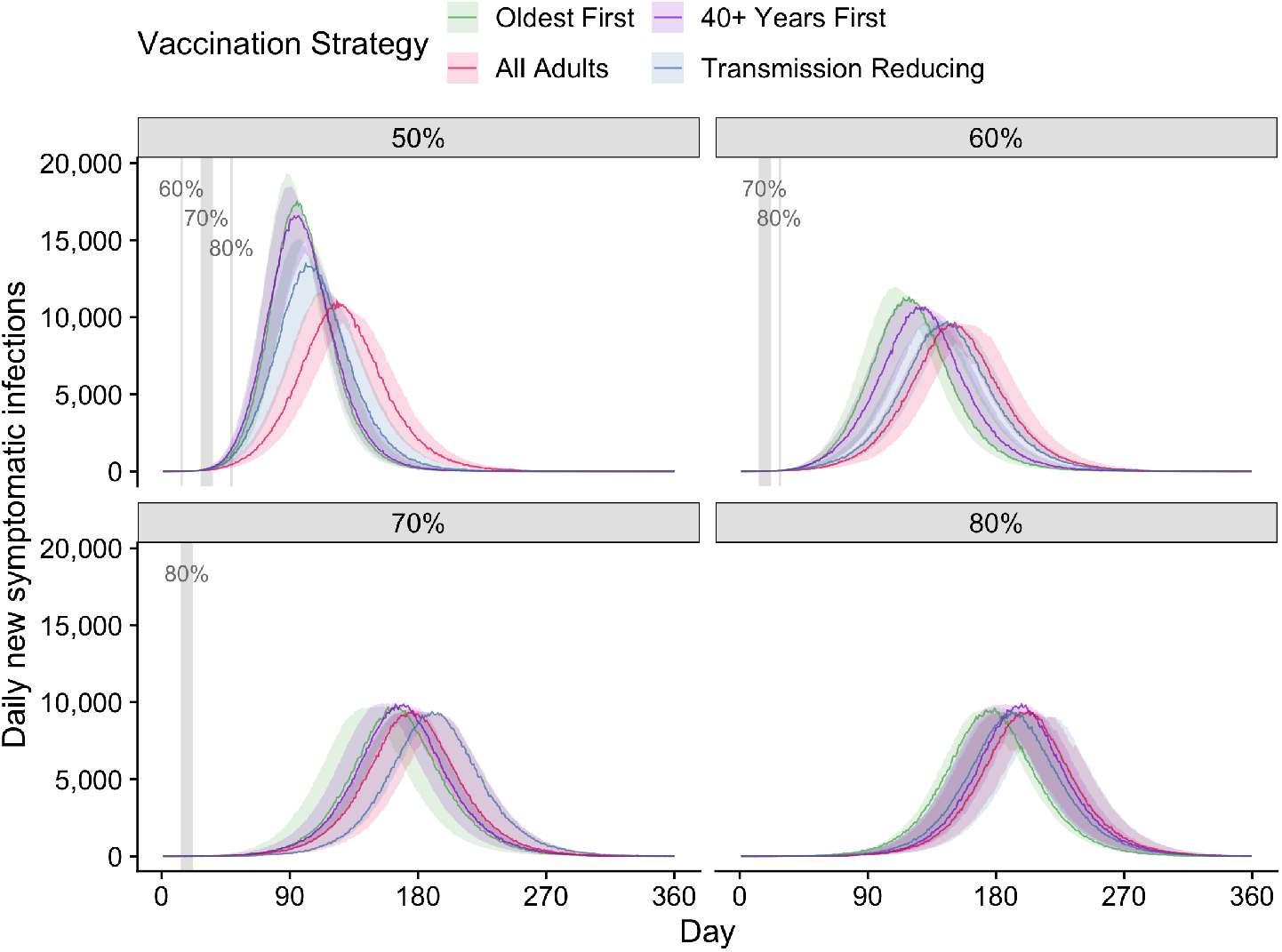
All four vaccination strategies — symptomatic infections. Simulations were seeded with a fixed, low number of infections (30 unvaccinated people).

**Figure S7.**
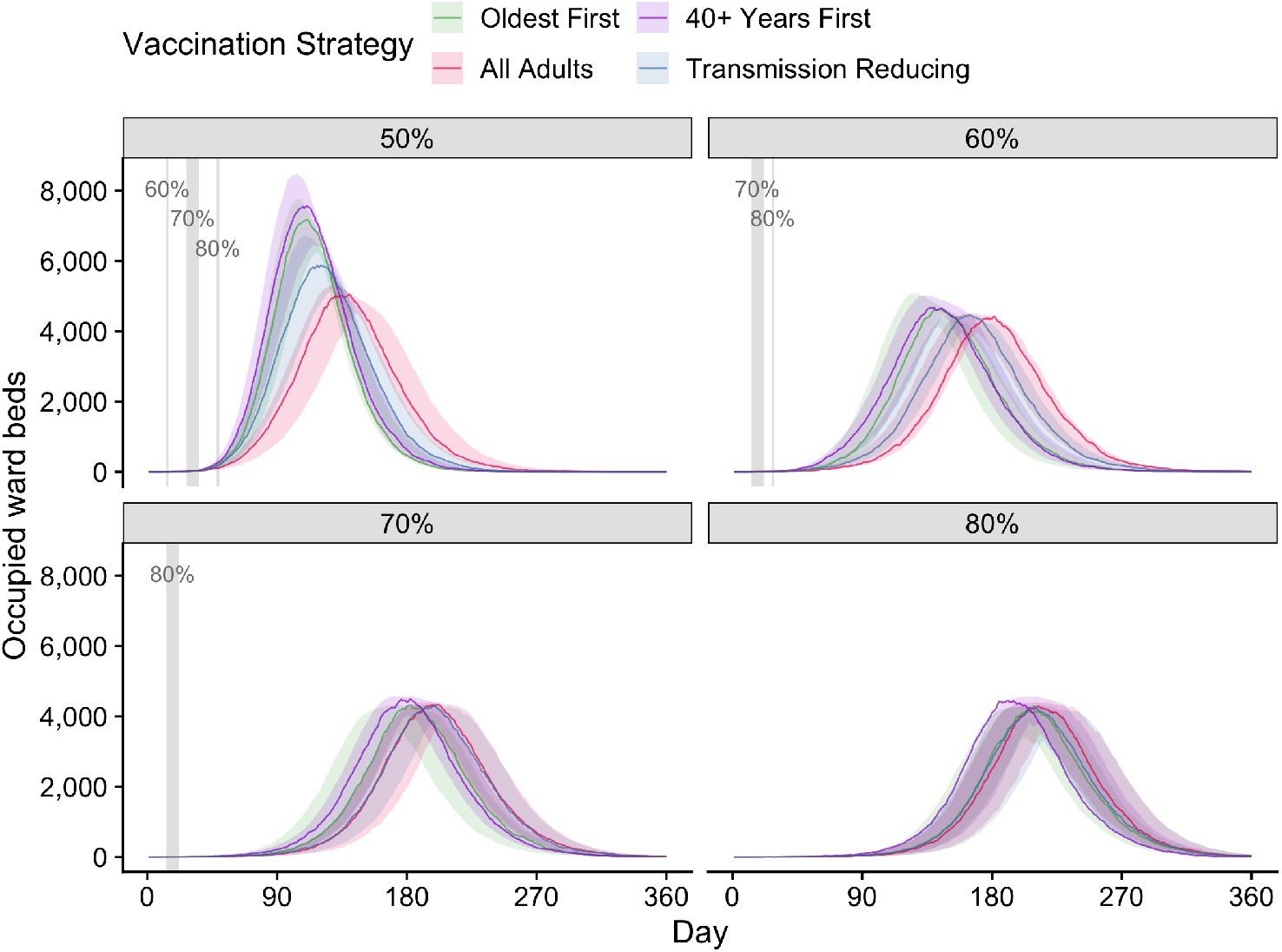
All four vaccination strategies — ward occupancy. Simulations were seeded with a fixed, low number of infections (30 unvaccinated people).

**Figure S8.**
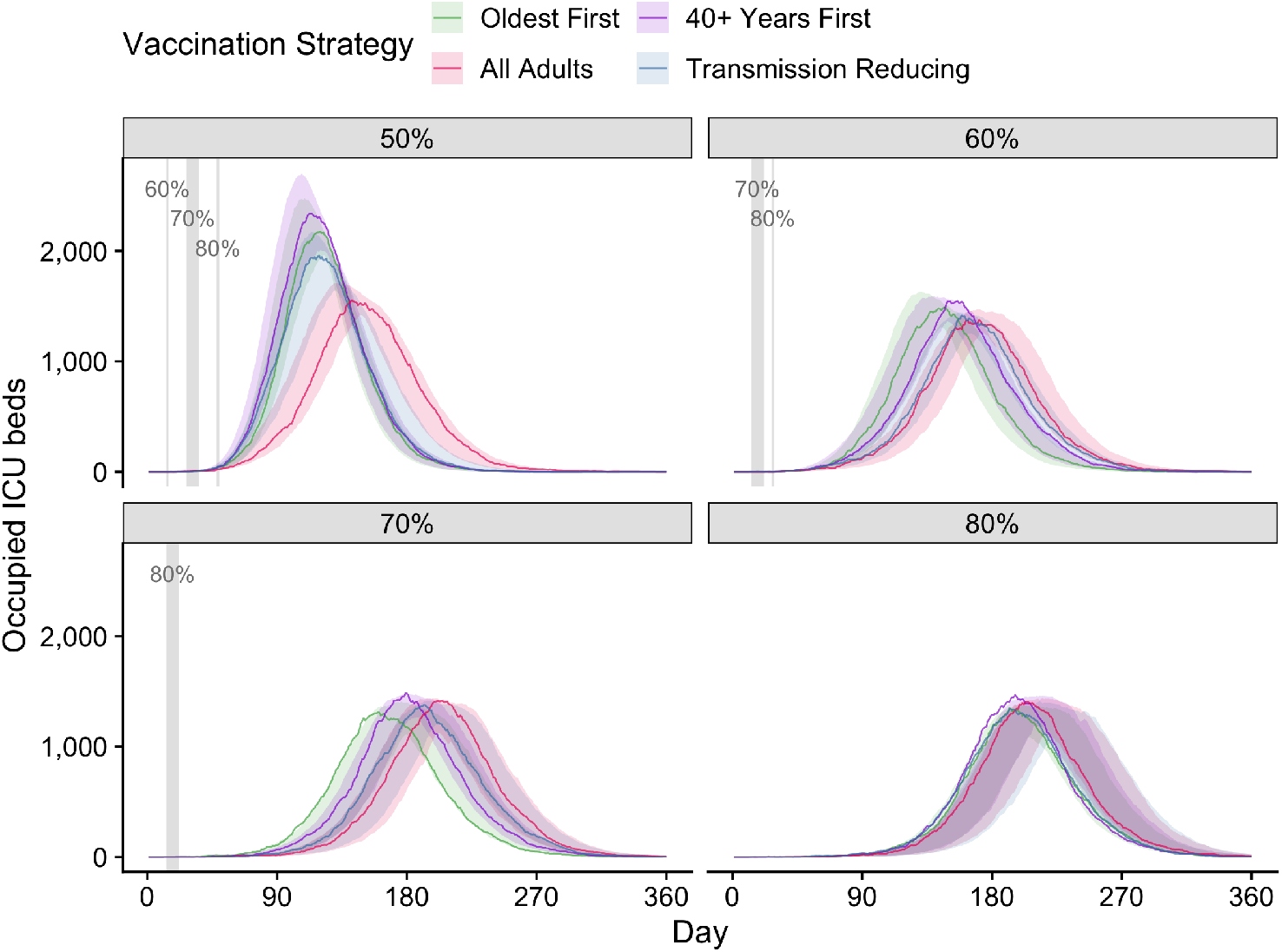
All four vaccination strategies — ICU occupancy. Simulations were seeded with a fixed, low number of infections (30 unvaccinated people).

**Figure S9.**
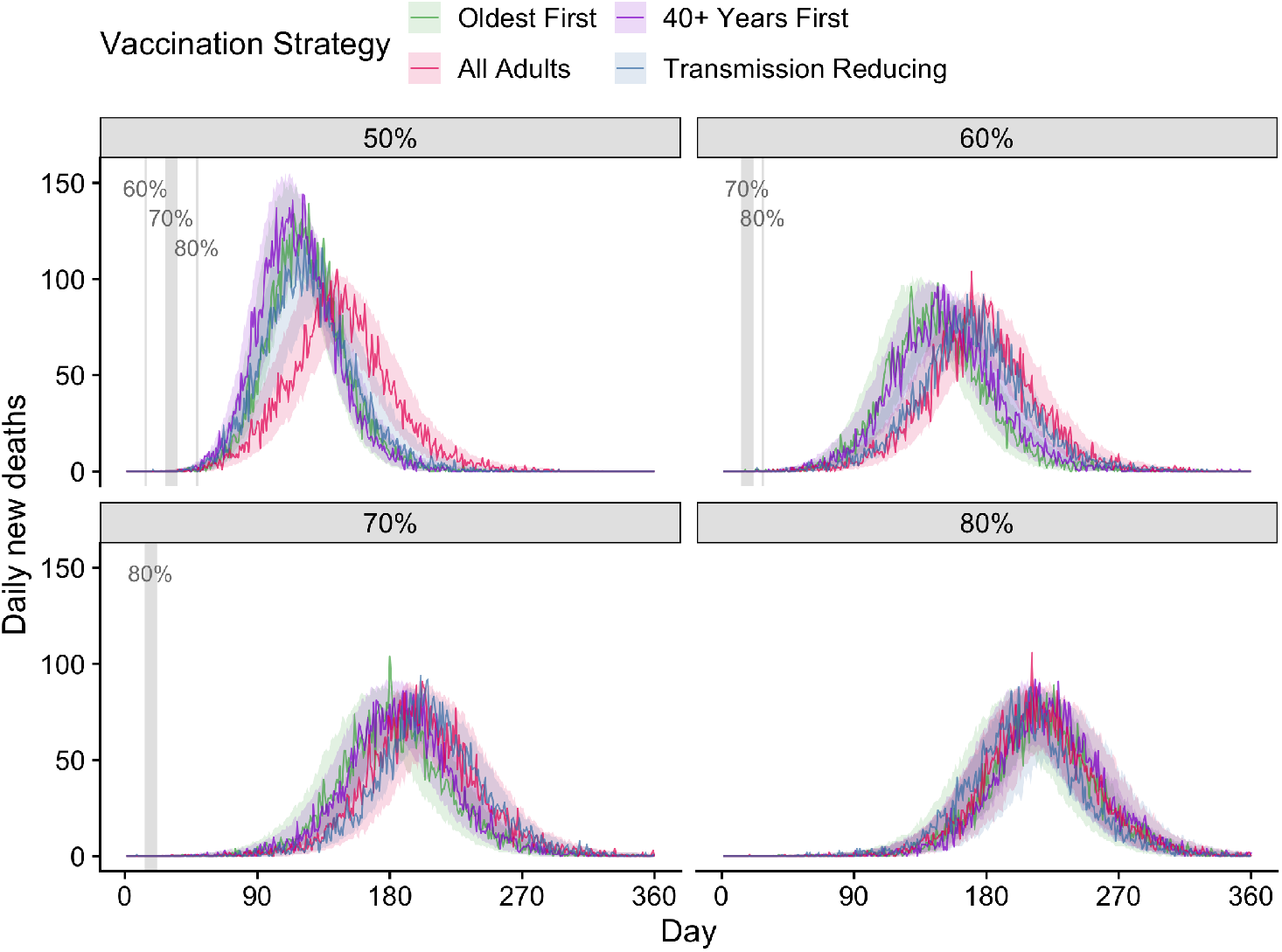
All four vaccination strategies — Deaths. Simulations were seeded with a fixed, low number of infections (30 unvaccinated people).

**Figure S10.**
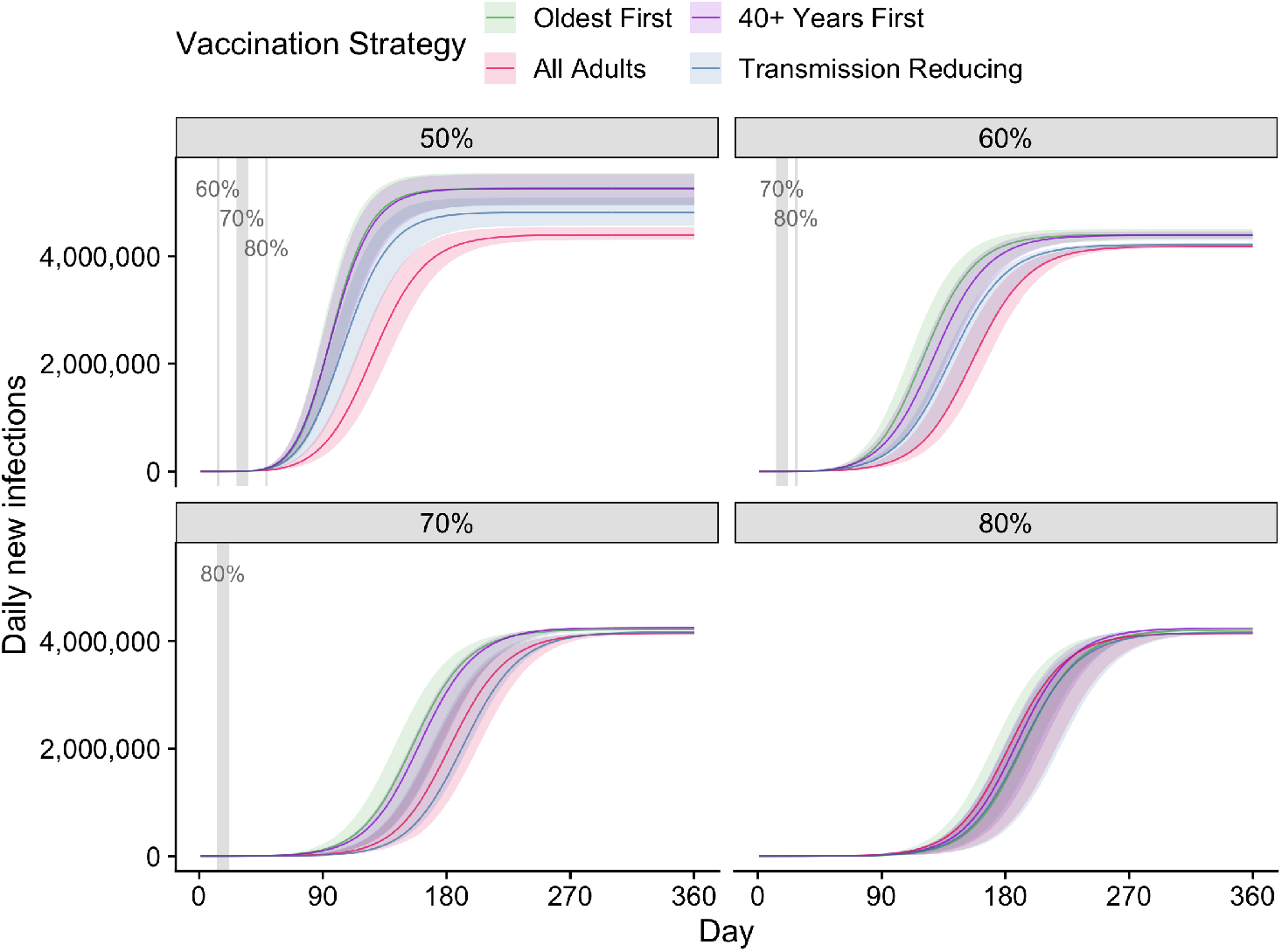
All four vaccination strategies — Cumulative infections (asymptomatic or symptomatic). Simulations were seeded with a fixed, low number of infections (30 unvaccinated people).

**Figure S11.**
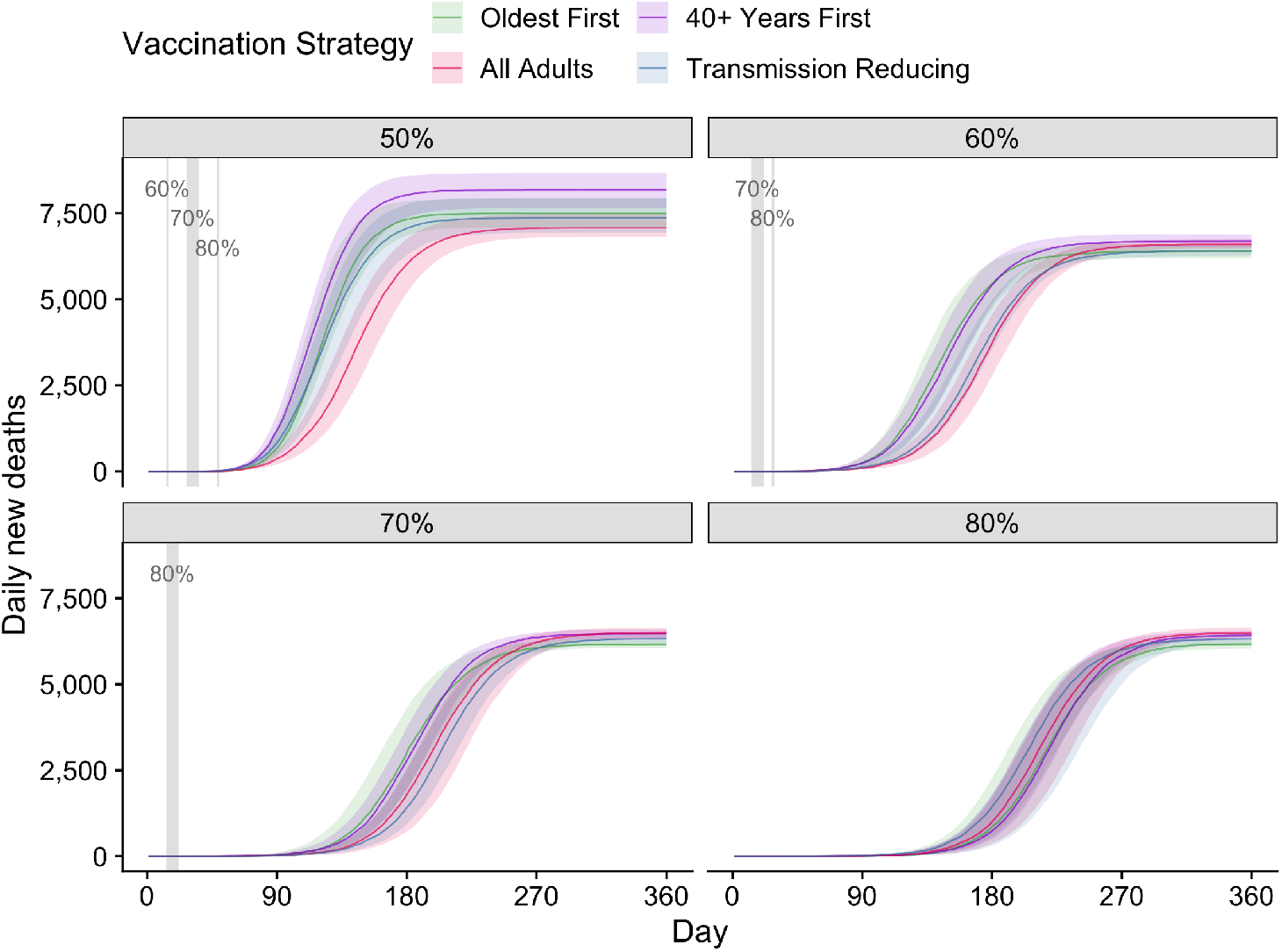
All four vaccination strategies — Cumulative deaths. Simulations were seeded with a fixed, low number of infections (30 unvaccinated people).

**Figure S12.**
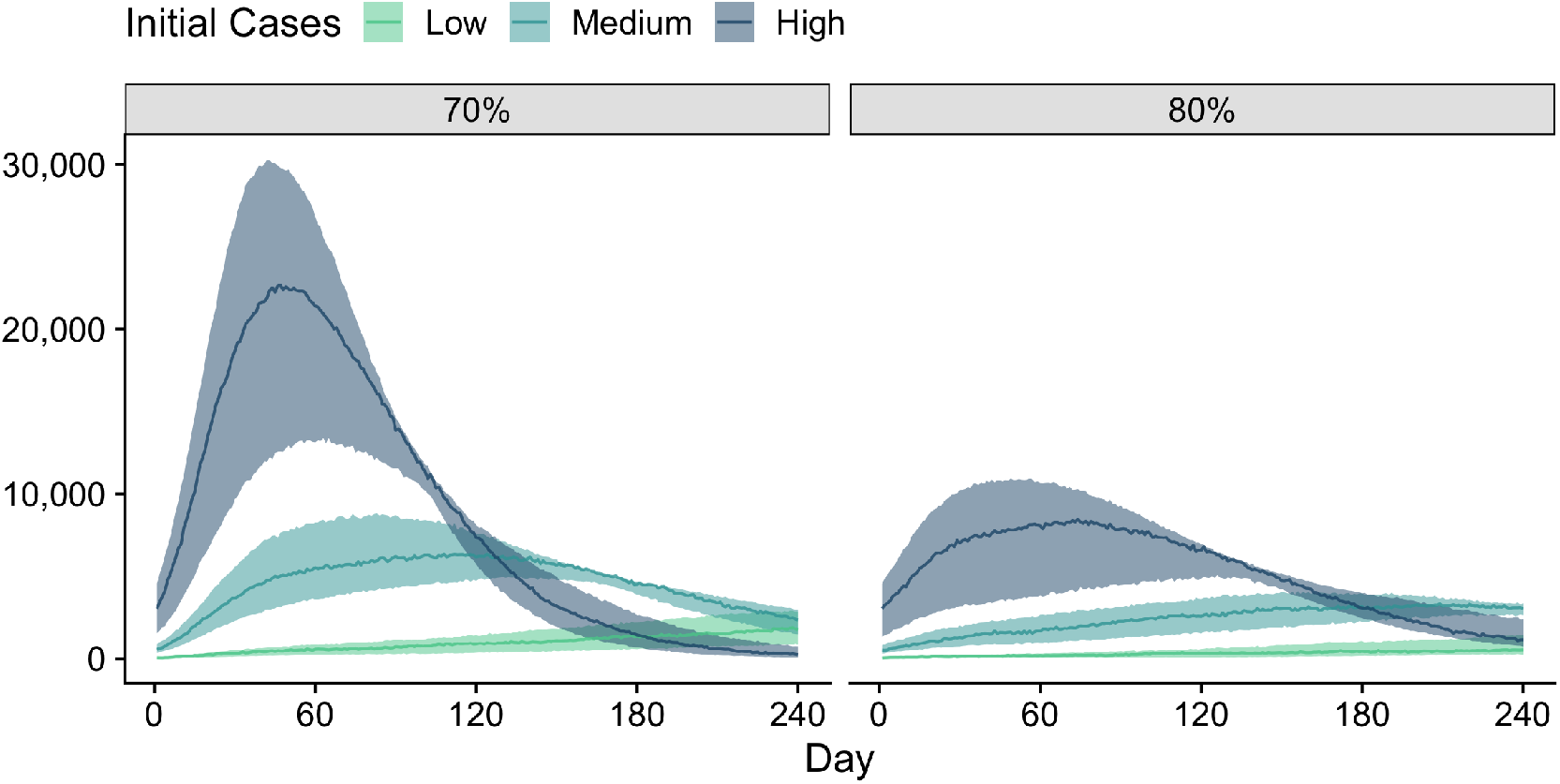
‘Low’, ‘Medium’ and ‘High’ initial infections at 70% and 80% vaccination coverage threshold with optimal TTIQ and baseline PHSMs.

**Figure S13.**
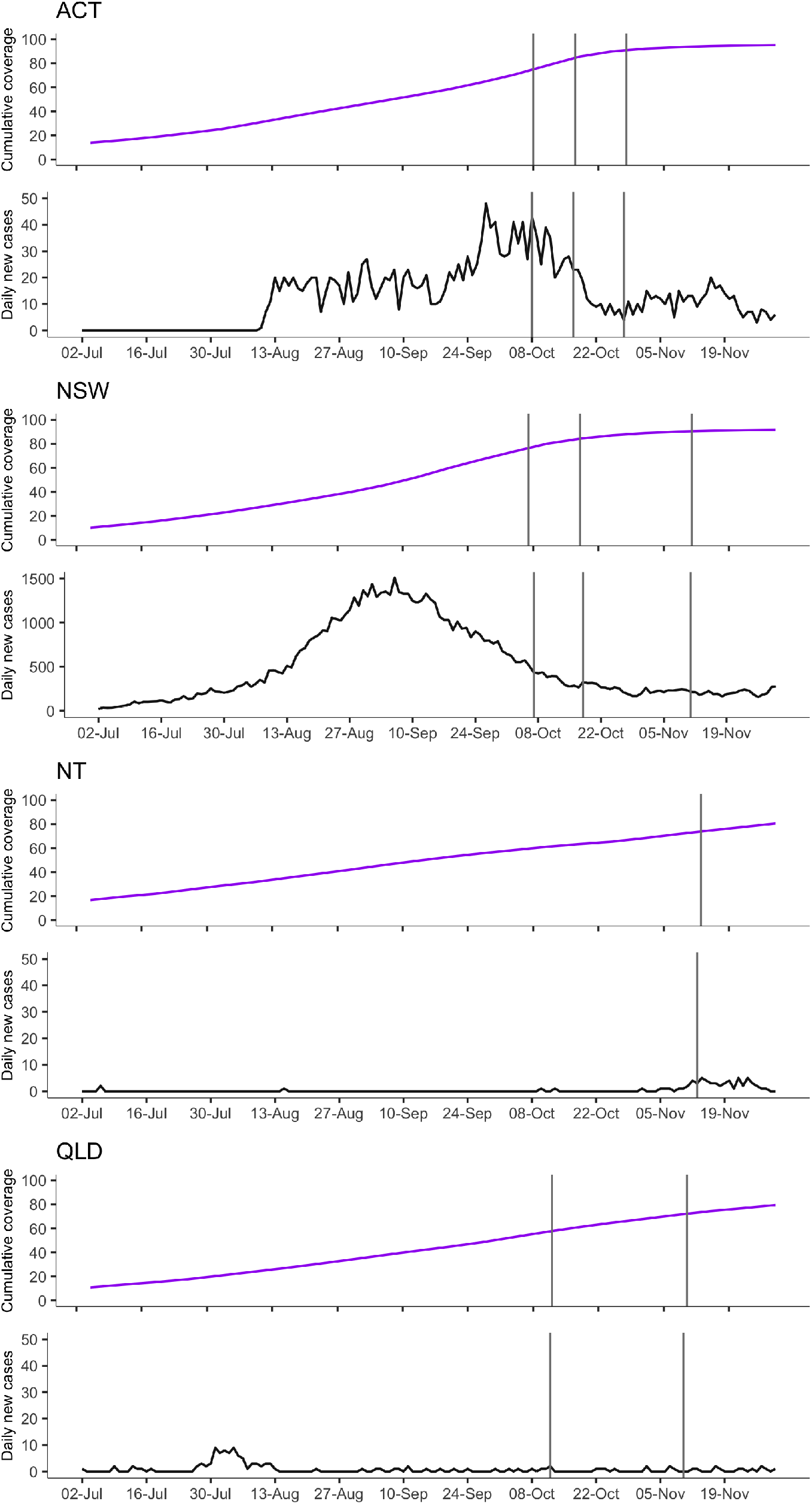
Time-series of cumulative vaccine coverage (purple lines) and daily case notifications (black lines) by Australian states/territories from 1 July 2021 up to 1 December 2021. Solid grey vertical lines (or shading) indicate the date of achieving the 70%, 80% and 90% vaccine coverage thresholds. ACT = Australian Capital Territory. NSW = New South Wales. NT = Northern Territory. QLD = Queensland.

**Figure S14.**
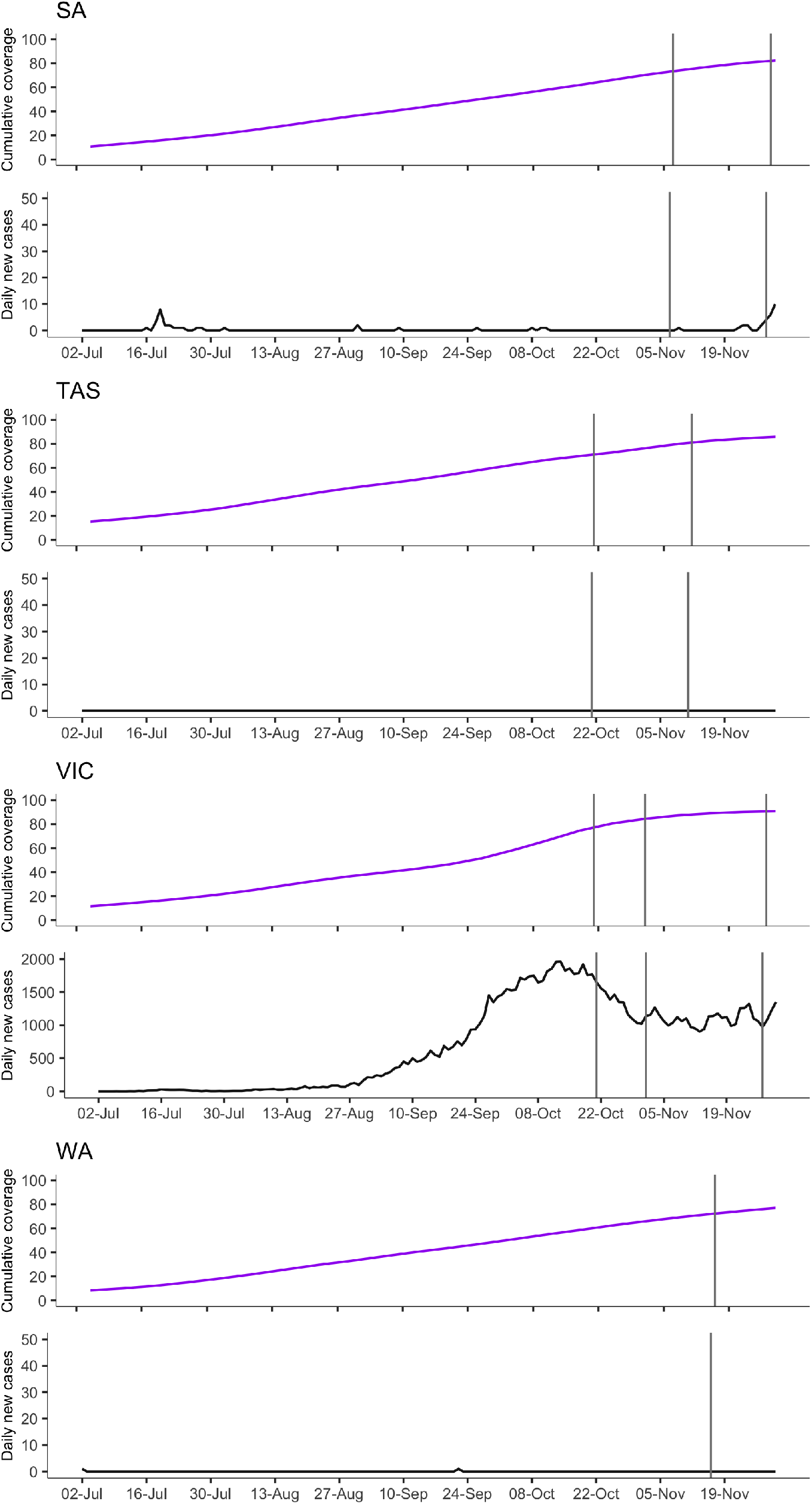
Time-series of cumulative vaccine coverage (purple lines) and daily case notifications (black lines) by Australian states/territories from 1 July 2021 up to 1 December 2021. Solid grey vertical lines (or shading) indicate the date of achieving the 70%, 80% and 90% vaccine coverage thresholds. SA = South Australia. TAS = Tasmania. VIC = Victoria. WA = Western Australia.

